# A knowledge database of physical activity interventions for hypertension using large language models

**DOI:** 10.64898/2026.10.22.26366246

**Authors:** Chunyu Li, Chikamara Obioha, Xingzhi Cui, Xiyan Zhou, Anthony Rosenzweig, Meng Wang

## Abstract

Physical exercise lowers blood pressure, but guidelines provide broad recommendations that rarely account for age, comorbidities, medications and hypertension severity. A structured, queryable resource of exercise interventions for hypertension is lacking, while manual curation is labor-intensive and difficult to scale. We present PAHD (Physical Activity for Hypertension Database), built with large language models (LLMs). An LLM screened 2,548 PubMed records through title-and-abstract and full-text assessment, selecting 255 studies. LLMs extracted arms, exercise protocols, participant characteristics, and outcomes, with two LLMs cross-checking study structure and each value linked to a verbatim quote. Programmatic integration generated 17,273 standardized records from 253 studies, one per reported comparison. Against adjudicated benchmarks, screening achieved 86.3% recall and 80.6% precision, and extraction 90.1% recall and 90.5% precision for blood pressure values. A clinician-facing portal retrieves studies matched to patients’ hypertension stage and comorbidities. PAHD is a reusable, traceable evidence resource for personalized exercise interventions in hypertension.

**Author summary:** Regular exercise lowers blood pressure, but people with high blood pressure differ in age, other illnesses and medications, and current guidelines give mostly general advice. Hundreds of clinical trials have tested specific exercise programs in specific groups of patients, but their findings are scattered across papers and laborious to collect by hand. We used large language models to find these trials and read them. The models selected 255 trials from 2,548 candidate records and recorded who participated, what exercise they did and how their blood pressure changed, linking every value to a quotation from the paper so that it can be checked. Compared with careful human reviews, the models captured most information accurately. The resulting database holds 17,273 records from 253 trials, and a website lets clinicians find the exercise programs that were tested in patients with characteristics similar to their own patients. We hope this resource helps researchers and clinicians use the existing trial evidence on exercise for hypertension.

## Introduction

Hypertension is characterized by persistently high blood pressure in the systemic arteries[1]. Blood pressure (BP) is recorded with two values: systolic BP, the pressure exerted on artery walls when the heart contracts, and diastolic BP, the pressure when the heart relaxes between beats[2]. Under the American College of Cardiology (ACC) / American Heart Association (AHA) guidelines[3], an individual is considered hypertensive when systolic BP is at least 130 mm Hg or diastolic BP is at least 80 mm Hg. These thresholds classify nearly half of US adults as hypertensive[4], underscoring its importance as a public health problem. The economic burden is substantial: projections from 2010 to 2030 estimate that total direct costs attributed to hypertension will nearly triple (from US $130.7 billion to US $389.9 billion) and indirect costs from lost productivity will almost double (from US $25.4 billion to US $42.8 billion)[5]. Effective prevention and treatment of hypertension are therefore critical to reducing disease burden and improving population health.

Physical activity (PA) is a recommended non-pharmacological intervention to control BP[1,5]. Numerous studies have demonstrated that regular PA improves cardiovascular fitness, lowers BP, and enhances quality of life[5–7]. Current guidelines on PA and hypertension management[3,8,9] are designed for general populations. For instance, CDC guidelines advise at least 150 minutes per week of moderate-intensity aerobic activity, plus muscle-strengthening activity on at least 2 days per week[9], while the American College of Sports Medicine (ACSM) suggests 30 minutes per day of continuous or accumulated exercise[8]. These guidelines, while valuable, are largely generic: even recent consensus advice that tailors the type of exercise to the blood pressure level[10] rarely accounts for individual-level factors such as age, comorbidities, or medication use, all of which shape the safety and efficacy of an exercise protocol.

Safe and effective exercise prescription requires tailoring protocols to individuals. A promising solution is to collect effective PA interventions for subpopulations, characterized by their individual-level attributes, into a structured, queryable knowledge base, which in turn requires a systematic and comprehensive review of the existing literature on PA interventions for hypertension. Prior efforts, such as PAHFKB for heart failure[11], highlight both the utility of such resources and the barriers to scaling them: paper screening and data extraction were performed manually, demanding extensive human effort. Given the growing volume of hypertension research and the need for continuous updates, such manual approaches are not sustainable.

Large language models (LLMs) make such automation feasible. They can answer free-text questions and extract information from documents without being trained for the specific task[12], and they have been used to turn scientific papers into large databases of structured records[13,14]. With careful task design, prompts and output schemas, they can take over much of the repetitive work of reading hundreds or thousands of papers, which makes it possible to build, and keep updating, a structured and personalized knowledge base of exercise interventions for hypertension with far less human labor. Recent work applies LLMs to the steps of evidence synthesis. In a study that each used ten published articles, Claude 2 extracted pre-specified data elements with an accuracy of 96.3%[15], and in six ongoing systematic reviews, LLM-assisted extraction was at least as accurate as extraction by humans alone[16]. The agreement of GPT-4 screening with human reviewers ranged from none to moderate, depending on the review stage and the language, but it was almost perfect for full texts with highly reliable prompts[17]. TrialMind links study search, screening and the extraction of clinical outcomes for meta-analysis[18]. Most of these tools automate one step of one review, and few evaluations keep a test set apart from prompt development[19,20]. However, these approaches have not established a reusable resource that keeps arm-level exercise protocols, baseline characteristics and results, each with a source quote. Such a resource is essential for ensuring evidence traceability, enabling comparisons across heterogeneous studies, and identifying the exercise protocols that were tested in patients with similar clinical characteristics. Meta-analyses of exercise and BP likewise rank exercise modes by their average effect[21]; they cannot show which protocol suits a patient with a given stage, comorbidity or medication.

To the best of our knowledge, no structured database currently exists for exercise interventions specific to hypertension. In this work, we fill that gap. Our contributions are: (i) a two-step LLM screening procedure (title and abstract, then full text); (ii) an evidence-grounded extraction procedure in six steps, in which two LLMs cross-check the structure of each study and every extracted value carries a verbatim source quote; (iii) PAHD, a standardized database of 253 studies with one record per reported comparison; and (iv) a clinician-facing web portal for profile-specific evidence retrieval. We evaluated screening and extraction against two benchmarks, each annotated independently by two annotators and adjudicated, and the portal with automated tests of its usability, reliability and summary faithfulness.

## Results

### Pipeline overview

Our PAHD construction has four stages: screening, information extraction, database construction, and a web portal (Fig 1). Large language models (LLMs) screen the studies and extract their content; a programmatic pipeline then builds the database without further LLM calls. In the screening stage, we retrieved candidate studies from PubMed with a Boolean query designed to miss as few relevant studies as possible. An LLM (OpenAI gpt-6-luna[22]) then judged each study against five eligibility criteria in two steps: first on its title and abstract, and then, for the studies that were not excluded on their title and abstract, on its full text.

**Fig 1.** Overview of the PAHD construction pipeline. a, Summary of the four stages of the pipeline (screening, information extraction, database construction and web portal) and their outputs. b, Detailed workflow. Screening: candidate studies retrieved from PubMed were screened by an LLM first on the title and abstract and then on the full text against the eligibility criteria. Information extraction: the skeleton and outcomes steps define the structure of each study (arms, interventions, outcomes and time points); two LLMs extracted it independently, and their disagreements were resolved by blinded cross-checking and adjudication. The study, interventions, arms and results steps then extract the remaining details, guided by the data schema, and every extracted value is paired with a verbatim quote from the source. Database construction: deterministic steps build one table per entity, standardize vocabularies and units, join the tables into one row per Result and check their quality, yielding PAHD. Portal: the web portal matches a patient profile (hypertension stage, comorbidities or preferred exercise) to studies in PAHD and returns the matched studies with their BP results, protocols, p-values and journals, with an optional AI summary. Icons mark LLM steps and deterministic (code) steps. AI, artificial intelligence; BP, blood pressure; FITT, frequency, intensity, time and type; LLM, large language model.

In the information extraction stage, six steps each extract one part of every eligible study, and together they collect the data items that systematic reviews collect from each study, as listed in the Cochrane Handbook: its methods, participants, interventions, outcomes and results[23]. The skeleton step identifies the arms, that is, the groups of participants that a study compares, and the interventions they receive. The outcomes step identifies the outcomes and the time points at which they were measured. Together, these two steps define the structure of a study, on which every later step depends. Two LLMs (gpt-6-luna and Google gemini-3.8-flash[24]) therefore extracted the structure independently. Where they disagreed, both reviewed the two answers blindly against the paper, and a third LLM (Anthropic Claude Sonnet 5.5[25]) adjudicated the disagreements that remained. The four other steps then fill in the details with gpt-6-luna: the study step extracts facts about the study as a whole, such as its design, region and sample size; the interventions step extracts the protocol of each intervention, with its frequency, intensity, session duration and type of exercise; the arms step extracts the baseline characteristics of each arm, such as age, sex, body mass index (BMI), medications and comorbidities; and the results step extracts every reported result of each outcome. Every extracted value is paired with a verbatim quote from the paper, so that it can be checked against the source.

In the database construction stage, four programmatic steps turn the extractions into the database. The tables step stores them in linked tables, one per entity of the data model (Fig 2): papers, arms, interventions, time points, outcomes, baseline statistics and results, connected by the identifiers that the skeleton and outcomes steps assigned. The standardization step removes results that cannot be used, such as comparisons between two baseline values, and maps free-text values to standard categories and units. The joining step combines the tables into one table, and the quality control step checks every record for internal consistency, corrects clear errors by rule and flags the rest. Each record of the final table is one comparison reported in a paper: an outcome, the two sides being compared (groups, time points or both), the statistics of each side and the difference between them. These records form PAHD.

**Fig 2.** Entity-relationship model of PAHD. A Paper holds study-level facts and is linked to the study structure through the paper identifier. Within the study structure, link tables record which arms receive which interventions and at which time points each outcome was measured. A Result is one comparison between two sides. For each side, it refers to rows of the Arm, Intervention and Time point tables and holds the statistic, its dispersion and the number analyzed. For the comparison, it refers to the Outcome and holds the difference between the sides, its CI and the p-value; each Result is one record of the database. Arrows mark foreign keys referencing a primary key. Only some fields of each table are shown. BMI, body mass index; BP, blood pressure; CI, confidence interval; FK, foreign key; PK, primary key; PMID, PubMed identifier.

Finally, a web portal lets clinicians enter a patient’s hypertension stage and either the patient’s comorbidities or a preferred type of exercise, and retrieve matching studies with their exercise protocols and BP results.

### Corpus construction and study selection

The identification and selection of candidate studies are summarized in the PRISMA flow diagram[26] (Fig A in S1 Appendix). The PubMed query returned 24,568 records within the publication period, of which 2,548 entered LLM screening after the removal of records without access to the abstract and full text, non-English or non-human records and records excluded for other reasons.

In the title-and-abstract step, the LLM judged each study from its title and abstract against five eligibility criteria: (1) the population included people with hypertension or elevated BP; (2) a structured exercise training program with multiple sessions was evaluated; (3) the effect of exercise could be isolated from other components of the intervention, such as diet; (4) the study was a completed intervention study with reported results; and (5) BP was reported as an outcome, with a statistically supported comparison involving an exercise arm. For each criterion, the LLM answered “Yes,” “No,” or “Unclear” (Methods). Studies with at least one “No” were excluded (2,087 removed). Full texts could not be retrieved for 8 of the remaining 461 studies, so 453 studies proceeded to full-text screening. In the full-text step, the LLM re-judged the same five criteria on the full text, accepting only “Yes” or “No.” It excluded 198 studies, most often because they did not study a hypertensive or elevated-BP population (102; Fig A and Note A in S1 Appendix). After screening, 255 studies were eligible for information extraction.

### Information extraction

All 255 eligible studies went through the six extraction steps. Each request to an LLM contained one whole paper: its text, converted from PDF to Markdown so that tables keep their rows and columns, and its figures as images, so that values shown only in a figure could also be read by the LLMs, all of which accept images as input. The prompts were developed on 14 of the included studies, which were kept out of the test set of the extraction benchmark. The two LLMs that extracted the structure of each study rarely disagreed. On the arms and interventions (skeleton step), they disagreed on at least one item in 23 of the 255 papers, and on the outcomes and time points (outcomes step) in 115; these disagreements were resolved by a blinded review by both LLMs or by the adjudicating LLM (Methods). Across all steps, the pipeline produced 53,782 evidence quotes. A programmatic check found 98.9% of the quotes taken from the text in the paper itself; the others were read from figure images (Note B in S1 Appendix). Screening and extraction took 10,183 LLM calls, which cost US$58.63 (US$0.23 per eligible study). A single call took on average 3 seconds in screening and 4–56 seconds in extraction. Calls for different papers can run in parallel (Table B in S1 Appendix).

### Screening accuracy

We measured screening accuracy on a benchmark of 500 of the 2,548 screened records (Table 1; Methods). Because eligible studies are rare, the sample was stratified: records whose title, abstract or MeSH terms mention both hypertension and exercise were sampled more heavily, and every record was weighted by the inverse of its stratum’s sampling rate, so that all estimates below describe the full pool of 2,548 records. Two annotators independently judged each record against the same five criteria, without seeing the pipeline’s decisions. They agreed on the eligibility decision for 98.4% of records (Cohen’s κ = 0.85; Gwet’s AC1 = 0.98)[27,28]. A human adjudicator then resolved, against the paper, every record on which the annotators’ answers disagreed (120 records), and this adjudicated label is the reference. After adjudication, 59 of the 500 records were eligible.

**Table 1.** Screening and extraction benchmarks.

| Stratum | Records in pool | Sampled | Eligible | Stratum weight |
|---|---|---|---|---|
| **a Screening benchmark** |  |  |  |  |
| S1 | 880 | 225 | 54 | 3.91 |
| S2 | 920 | 200 | 5 | 4.60 |
| S3 | 748 | 75 | 0 | 9.97 |
| **Total** | 2,548 | 500 | 59 | – |
| **b Extraction benchmark** |  |  |  |  |
| **Benchmark records** |  | **All** | **Test** | **Development** |
| Studies |  | 49 | 36 | 13 |
| Arms |  | 118 | 82 | 36 |
| Interventions |  | 118 | 82 | 36 |
| Time points |  | 125 | 93 | 32 |
| Outcomes |  | 902 | 596 | 306 |
| Baseline statistics |  | 803 | 574 | 229 |
| BP result rows |  | 988 | 639 | 349 |
| BP data points |  | 1,211 | 868 | 343 |

Each benchmark was annotated independently by two annotators, and every disagreement was adjudicated against the paper. **a**, Stratified sample of the 2,548 records that entered screening. Strata were defined by keyword flags in the title, abstract and MeSH terms: S1, hypertension and exercise terms; S2, exercise term only; S3, no exercise term. The stratum weight is the inverse of the stratum’s sampling fraction. Eligibility is after adjudication; 380 labels were agreed by both annotators and 120 were adjudicated. **b**, Fifty studies drawn from the included set, of which one meta-analysis was excluded. The development set was used during prompt development; the test set was held out. Result rows and data points cover blood pressure outcomes only. BP, blood pressure.

We report recall (the proportion of eligible studies that the pipeline kept), precision (the proportion of the studies it kept that were eligible), specificity (the proportion of ineligible studies that it excluded) and the F2 score, which combines recall and precision but weighs recall more heavily, because a study that is missed at screening is lost for good, whereas a wrongly kept study can still be removed later. A study counted as kept only if it passed both screening steps.

Screening was both sensitive and specific (Table 2; Fig 3a). The pipeline kept 86.3% of eligible studies (95% confidence interval [CI] 76.8–94.5%), and 80.6% of the studies it kept were eligible (70.6–90.5%). Specificity was 97.9% (96.6–99.1%), and the F2 score was 85.1% (77.0–91.9%). The title-and-abstract step alone kept 91.7% of eligible studies. In total, the pipeline passed on an estimated 251 of the 2,548 records.

**Fig 3.** Accuracy of screening and extraction. a, Screening accuracy against the 500-record benchmark, for the end-to-end decision (both screening steps) and for the title-and-abstract step alone; estimates are weighted to the 2,548 screened records. b, Extraction accuracy on the 36 test studies, grouped by the extraction steps that produce the fields. Points are estimates and lines are 95% CIs (a, stratified bootstrap of records; b, bootstrap of papers with 2,000 replicates). The FITT parameters were scored with an LLM judge for pairs that deterministic rules could not settle. BP, blood pressure; CI, confidence interval; F1, harmonic mean of precision and recall; F2, F-score weighting recall more heavily; FITT, frequency, intensity, time and type.

**Table 2.** Screening performance against the benchmark (weighted).

| **Metric** | **End to end** | **Title and abstract alone** | **Full text†** |
|---|---|---|---|
| Recall (sensitivity) | 86.3 (76.8–94.5) | 91.7 (83.8–98.2) | 94.2 (87.8–100.0) |
| Precision | 80.6 (70.6–90.5) | 46.4 (37.8–55.5) | 80.6 (70.9–89.9) |
| F1 score | 83.4 (75.6–90.3) | 61.6 (53.0–69.7) | 86.9 (80.3–92.6) |
| F2 score | 85.1 (77.0–91.9) | 76.7 (69.0–82.9) | 91.1 (85.5–95.8) |
| Specificity | 97.9 (96.6–99.1) | 89.3 (86.7–91.9) | 80.4 (69.9–90.1) |

Values are percentages with 95% confidence intervals in parentheses. Estimates are weighted to the screening pool of 2,548 records by the inverse of each stratum’s sampling fraction; intervals come from a stratified bootstrap of studies. The end-to-end decision counts a study as included only if it passed both screening steps.
†Evaluated on the 109 benchmark studies that passed title-and-abstract screening.

The pipeline missed 8 of the 59 eligible benchmark studies and wrongly kept 11 ineligible ones. Seven of the eight missed studies were dropped on criterion 5 (a statistically supported BP comparison), and all 11 wrongly kept studies failed the population criterion, mostly because they enrolled broader groups of whom only some had hypertension. Errors concentrated on studies that were hard for the annotators as well: 17 of the 19 errors concerned studies on which the two annotators had disagreed and the adjudicator decided (Note C in S1 Appendix).

### Extraction accuracy

We evaluated extraction against a benchmark of 49 of the 255 included studies (Table 1; Methods), drawn from the included studies by type of exercise so that every type is represented. For each study, two annotators independently recorded the same fields that the pipeline extracts, using the same field definitions, and their records were aligned field by field. A human adjudicator then checked every field on which they disagreed against the paper and set its final value; fields on which they agreed were kept as they were. 13 of the 49 studies had been used to develop the prompts (development set). The other 36 were not used in prompt development and form the test set, on which we report all results below, with 95% CIs.

We used three kinds of measures. For items that the pipeline has to find in a paper, such as arms, outcomes or BP values, we report recall, precision and the F1 score. For fields that take one value per study, arm or intervention, such as the design or the number of participants randomized, we report accuracy, the proportion of values equal to the benchmark value; a value that belongs to an arm or intervention that the pipeline missed counts as wrong. For fields with a fixed list of categories, we also report macro-F1, the average of the F1 scores of the categories. Confidence intervals come from resampling whole papers with replacement (bootstrap)[29].

The structure of each study was recovered almost perfectly (Table 3; Fig 3b). Arms, interventions and the links between them matched the benchmark in every test study (F1 = 100%). Time points reached an F1 of 97.3% (92.4–100.0%), all outcomes 94.0% (90.7–97.0%) and BP outcomes 99.4% (98.6–100.0%). Compared with the structure that each LLM extracted on its own, cross-checking mainly helped the arms and the BP outcomes (Note D in S1 Appendix).

**Table 3.** Extraction accuracy on the held-out test set.

| **Measure** | **n** | **% (95% CI)** |
|---|---|---|
| *Study structure (skeleton and outcomes)* |  |  |
| Arms, F1 | 82 | 100.0 (100.0–100.0) |
| Time points, F1 | 93 | 97.3 (92.4–100.0) |
| Outcomes, F1 | 596 | 94.0 (90.7–97.0) |
| BP outcomes, F1 | 177 | 99.4 (98.6–100.0) |
| *Study and arm facts (study and arms)* |  |  |
| Design, accuracy | 36 | 100.0 (100.0–100.0) |
| Total randomized, accuracy | 36 | 97.2 (91.7–100.0) |
| Assigned per arm, accuracy | 82 | 93.9 (84.5–100.0) |
| Completed per arm, accuracy | 82 | 79.3 (64.3–92.3) |
| Baseline statistics, F1 | 574 | 84.3 (78.7–89.2) |
| *Interventions* |  |  |
| Intervention type, accuracy | 82 | 87.8 (79.5–95.2) |
| Framed as control, accuracy | 82 | 96.3 (92.4–100.0) |
| Supervision, accuracy | 82 | 80.5 (69.8–90.6) |
| FITT parameters, accuracy† | 410 | 81.2 (76.5–85.1) |
| Protocol sentences recovered‡ | 273 | 90.5 (82.9–96.0) |
| Extracted protocol sentences found verbatim | 442 | 99.8 (99.3–100.0) |
| *Blood pressure results* |  |  |
| Data points, precision | 864 | 90.5 (83.7–95.5) |
| Data points, recall | 868 | 90.1 (84.1–94.4) |
| Data points, F1 | 868 | 90.3 (84.3–94.6) |
| Meta-analysis-ready data points§ | 868 | 77.8 (69.3–86.4) |
| Extracted values printed in the paper | 864 | 99.1 (97.2–100.0) |

Test set of 36 studies not used during prompt development. n is the number of benchmark items, or of extracted items for precision, protocol fidelity and printed values. Values are percentages with 95% confidence intervals from a paper-level cluster bootstrap (2,000 replicates). Accuracy is end-to-end: a value of a missed entity counts as wrong.
†Frequency, session duration, intensity, modality and total duration. Pairs not settled by deterministic rules were judged by an LLM; counting every judged pair as wrong or as correct gives bounds of 70.2% and 93.2%.
‡Benchmark protocol sentences that appear verbatim in the paper.
§Value and dispersion both correct, so that an effect size can be computed without returning to the paper.
BP, blood pressure; FITT, frequency, intensity, time and type; LLM, large language model.

Facts about each study were mostly accurate. The study design (for example, a parallel-group or crossover randomized trial) was correct for all 36 test studies, the total number of participants randomized for 97.2%, the number assigned to each arm for 93.9% and the number completing each arm for 79.3%. The type of each intervention, one of ten categories such as exercise, exercise combined with another component, usual care or no intervention, was correct for 87.8% (79.5–95.2%) of interventions. Whether the paper framed an intervention as a control was correct for 96.3% of interventions, and the level of supervision for 80.5%.

Baseline characteristics were mostly captured. A baseline statistic is one reported value of one characteristic for one arm, such as the mean age of the exercise arm with its standard deviation, or the number of women in the control arm. The pipeline found 87.8% of the benchmark’s baseline statistics (recall), and 81.0% of the statistics it extracted matched a benchmark statistic (precision), for an F1 of 84.3% (78.7–89.2%).

The exercise prescription of each intervention was usually captured (Table 3; Fig 3b). For each intervention, the pipeline copies the verbatim protocol description and extracts five prescription parameters as short texts: frequency, session duration, intensity and modality (the FITT components) and the total duration of the program. To score the content, we compared each of the 410 extracted parameters (82 test interventions × 5 parameters) with the benchmark text. A pair counted as correct when the two texts were the same after normalization, when the extraction contained the benchmark text, or when their numbers and units matched; the remaining pairs were judged by a second LLM (Methods). By this measure, 81.2% (76.5–85.1%) of the parameters were correct.

Frequency was the most accurate parameter (92.7%), followed by session duration (85.4%), modality (80.5%) and intensity (75.6%). Total program duration was the least accurate (72.0%), mostly because of a difference in convention for control arms that received no intervention (Note D in S1 Appendix).

The verbatim protocol descriptions were faithful to the paper: 99.8% of the sentences of the extracted descriptions were found in the paper text, and the extracted descriptions recovered 90.5% of the benchmark protocol sentences that appear in the paper (Table 3). Almost all of their text came from the Methods section (Note D in S1 Appendix).

A row of a results table usually holds several numbers: for example, the mean SBP of the exercise arm at baseline and after training, each with its standard deviation, and the difference between the arms with its p-value. We therefore scored the BP results number by number, as data points, rather than as table rows. A data point is the statistic of one group at one time point (such as the mean SBP of the exercise arm after 12 weeks), the change of one group between two time points, or one difference between groups. A data point counted as correct when its outcome, groups and time points matched the benchmark and its value was equal. The pipeline recovered 90.1% (84.1–94.4%) of the 868 benchmark BP data points, and 90.5% (83.7–95.5%) of the data points it extracted were correct, for an F1 of 90.3% (84.3–94.6%). For 77.8% (69.3–86.4%) of the benchmark data points, both the value and its dispersion (standard deviation, standard error or CI) were correct, so that the data point could enter a meta-analysis without returning to the paper. Values were rarely invented: 99.1% of the extracted BP values appear in the paper text.

We assigned each benchmark data point that was not fully correct to one error category (Fig 4). The most common error concerned the dispersion and affected 12.3% of all benchmark data points (107 of 868, 55% of all errors): the value was right, but its dispersion was missing or wrong. Most often (59 of these 107), the pipeline did not record the type of dispersion, typically because the paper stated it only once, in its statistical methods, rather than next to the numbers. These errors were concentrated: three papers accounted for 80 of the 107. Omissions, in which the pipeline did not extract a benchmark value at all, affected 4.4% of data points; values attributed to the wrong group or time point 2.4%; a reversed sign, such as a decrease recorded as an increase, 1.7%; and a wrongly copied value 1.4%. Among the data points that the pipeline matched, the value was correct for 95.2%, but the number of participants analyzed in the group was correct for only 53.9%, mostly because the pipeline left it empty (Note D in S1 Appendix).

**Fig 4.** Errors in the extraction of blood pressure results. a, Category of each of the 868 benchmark BP data points of the 36 test studies: correct (value and dispersion correct), dispersion error (value correct; dispersion missing or different), omission (value not extracted), misattribution (value extracted for another group, time point or outcome), sign error (value with the opposite sign) and transcription error (data point extracted with a different value). Bars show the proportion of the 868 data points with 95% CIs. b, Number of errors in each of the 26 test studies with at least one error; three studies account for 80 of the 107 dispersion errors. c, One example of each error category from the test set, with the value that the benchmark records and the value that the pipeline extracted. BP, blood pressure; CI, confidence interval; SD, standard deviation; SE, standard error.

### Database construction and overview

The database construction stage turned the extractions of the 255 studies into PAHD. During standardization, two studies were removed, one because none of its arms received exercise (it compared health education with usual care) and one because it reported BP only as correlations with other measures, resulting in 253 studies. Automated quality checks then tested every record for internal consistency, for example whether a reported difference matches the two group values and whether every number appears in the paper. Most records (89%) passed every check. Clear errors, such as a reversed sign, were corrected by rule, flagged BP records were reviewed by hand against the paper, and the remaining flags stay in the database so that users can treat those records with care (Table 4; Methods).

**Table 4.** Key columns of PAHD.

| Group | Column | Rows with a value (%) | Most common values (% of rows with a value) |
|---|---|---|---|
| Study | Study design | 100 | parallel RCT (89%); crossover RCT (4%); non-randomized (3%) |
|  | Region (ISO code) | 60 | BRA (21%); USA (12%); CHN (8%) |
| Comparison | Arms compared | 100 | exercise arm over time (41%); exercise vs control (27%); control arm over time (24%) |
|  | Comparison type | 100 | one arm over time (68%); groups, change (19%); groups, one time point (12%) |
| Result | Outcome | 100 | SBP (5%); DBP (5%); BMI (2%); Body weight (2%); any BP outcome (28%) |
|  | p-value | 71 | p < 0.05 in 52% of rows with a p-value |
| Exercise | PA Category | 76 | Aerobic (41%); Aerobic, Resistance (16%); Resistance (16%) |
|  | PA Frequency | 72 | 3 sessions/week (53%); 2 sessions/week (13%); 1 session/day (4%) |
|  | PA Session Duration | 66 | 60 minutes (24%); 30 minutes (7%); 40 minutes (4%) |
|  | PA Volume | 64 | median 120 (IQR 90–180) min/week (lower bound of a range) |
|  | PA Program Duration | 75 | 12 weeks (27%); 8 weeks (13%); 16 weeks (8%) |
|  | Supervision | 59 | fully supervised (64%); partly supervised (24%); unsupervised (5%) |
| Population | Age | 67 | median 58 (IQR 51–65) years (arm mean) |
|  | Sex | 47 | Female (85%); Male (79%) |
|  | Race and ethnicity | 8 | White (83%); Black or African American (57%); Asian (36%) |
|  | BMI category | 65 | Overweight (74%); Obesity (57%); Normal (24%) |
|  | Hypertension stage | 50 | Stage 2 (34%); Stage 1 (33%); Elevated (14%) |
|  | Medications | 34 | Calcium channel blockers (49%); Angiotensin II receptor blockers (48%); ACE inhibitors (47%) |
|  | Comorbidities | 20 | Diabetes mellitus (41%); Hypertension (34%); Dyslipidemia (23%) |
| Quality | Quality-control status | 100 | clean (89%); flagged (6%); reviewed (2%); auto-fixed (2%) |

Columns of the PAHD table (one row per reported comparison; 17,273 rows). The exercise and population columns describe the exercise (intervention) side of a row. Percentages are proportions of rows, not of studies, and a row can list several values (for example, several medication classes). Weekly volume is the lower bound of the prescribed minutes per week. Age is the central value of the arm. IQR, interquartile range; BP, blood pressure; RCT, randomized controlled trial.

PAHD contains 17,273 records from 253 studies; each record is one reported comparison. Table 4 lists its key columns, the proportion of records in which each column has a value and its most common values, and Fig 5 summarizes the contents of the database. The studies describe 623 arms, 619 interventions and 4,665 outcomes. Most were parallel-group randomized trials (228; 90%), and the studies included 22,776 participants in total (Note E in S1 Appendix).

**Fig 5.** Contents of PAHD. a, Studies by year of publication. b, Proportion of the 253 studies that report each characteristic of the study population. c, The 368 exercise interventions by type of exercise and by the most common standardized values of sessions per week, session duration and program length; an intervention can combine several types of exercise. d, Within-arm changes in SBP and DBP in exercise and control arms (mean differences in mm Hg, later minus earlier time point): median (dot), interquartile range (bar) and 10th to 90th percentile (line). BMI, body mass index; BP, blood pressure; DBP, diastolic blood pressure; SBP, systolic blood pressure.

The database records the arm-level characteristics that are needed to match studies to patients (Fig 5b). Age was reported in 99% of studies, sex in 97% and body mass index in 90%, and the median of the study mean ages was 58 years (interquartile range [IQR] 51–65). Baseline BP allowed a 2025 AHA/ACC stage[3] to be assigned in 62% of studies. Race and ethnicity were the most sparsely reported characteristics, present in only 22% of studies (Note E in S1 Appendix).

Clinical-context fields provide the medication and comorbidity detail. Medication use was reported in 51% of studies and comorbidities in 42%; diabetes was the most common comorbidity (25%; Note E in S1 Appendix).

We describe the exercise interventions by the four components of an exercise prescription: frequency, intensity, time and type (FITT)[30]. Of the 368 exercise interventions (Fig 5c), 63% included aerobic and 37% resistance training. Neuromotor exercise such as yoga or tai chi (5%), flexibility training (2%) and other forms such as breathing exercise or inspiratory muscle training (12%) were less common. Three sessions per week was by far the most common frequency (57%). The most common session durations were 60 minutes (22%) and 30 minutes (8%). Programs most often lasted 12 weeks (29%) or 8 weeks (17%). Note E in S1 Appendix summarizes their weekly volume, intensity and supervision.

The database is rich in quantified outcome evidence. Of the 17,273 records, 11,709 (68%) compare the same arm at two time points, such as before and after the intervention, and 5,342 (31%) compare two groups, either at the same time point or by their changes over time; the remaining 222 (1%) are other comparisons, for example between the conditions of a crossover trial. Classified by the arms involved, 7,084 records follow an exercise arm over time, 4,633 compare an exercise arm with a control arm, and 4,063 follow a control arm over time. BP outcomes account for 4,855 records (28%) from all 253 studies. Secondary outcomes include body weight and BMI, blood lipids, heart rate, glucose and fitness. The extracted effects agree with the known antihypertensive benefit of exercise: of 933 within-arm changes in systolic or diastolic BP in exercise arms, 82% were decreases, 14% increases and 4% showed no change.

### Web portal and its evaluation

The PAHD web portal lets a clinician to look up studies that fit a patient (Fig 6). The user enters the patient’s hypertension stage, or a BP reading from which the stage is derived, and then either the patient’s comorbidities or a preferred type of exercise. The portal returns up to five studies. Each is shown with its main BP result, the exercise protocol (frequency, intensity, session duration and modality), the p-value, the journal and, where available, the journal’s Journal Impact Factor (JIF) from Clarivate’s Journal Citation Reports[31]. The user can ask for a short AI-written summary of the protocols they select. The portal searches only results that involve an exercise arm and a BP level measured at rest, over 24 hours, at home or centrally (in the aorta): 3,117 comparisons from 248 of the 253 studies. The other five studies have no such result; for example, they report BP only during a tilt or exercise test, or only its variability. Note F in S1 Appendix gives the hypertension stages and comorbidities of the 248 studies.

**Fig 6.** The PAHD web portal. a, Query panel: the user chooses how many studies to show, enters the patient’s hypertension stage (or a BP reading from which the stage is derived) and chooses to match by comorbidity or by preferred type of exercise; comorbidities can be selected or typed in free text. b, Matched studies, with an explanation of how they were matched; each row shows the rank and type of match (exact or similar), the main BP result, the FITT parameters, the p-value and the JIF from Clarivate Journal Citation Reports. c, Details of one study: the verbatim protocol, its FITT parameters, the study population and the group statistics of the displayed result. d, Optional AI-written summary of the selected protocols, which ends with a disclosure that it is AI-generated. i and ii, The first row of the table in b, magnified (outlined in b). Numbered markers: 1, hypertension stage, or a BP reading from which the stage is derived; 2, matching by comorbidity or by preferred type of exercise; 3, comorbidities selected from the list or typed in free text; 4, plain-language explanation of how the studies were matched; 5, rank and type of match; 6, JIF; 7, verbatim protocol and its FITT parameters; 8, group statistics of the displayed BP result; 9, disclosure that the summary is AI-generated. BP, blood pressure; FITT, frequency, intensity, time and type; JIF, Journal Impact Factor.

We tested the portal with code, without human raters or an LLM judge, by calling the functions that the portal’s server uses (Table 5; Methods). The tests used 440 queries of three types: 320 queries with a single comorbidity (each of the 64 comorbidity options, including none, with each of the four stages and an unknown stage), 50 queries with random combinations of two or three comorbidities, and 70 queries by exercise type (each of the 14 exercise categories with each stage and an unknown stage). Each query asked for five studies. No query ended without a result, and a query took a median of 0.02 s (95th percentile 0.08 s).

**Table 5.** Automated evaluation of the web portal.

| Test | Result |
|---|---|
| *Usability* |  |
| Queries answered with five studies, by comorbidity | 370/370 (100%) |
| Queries answered with five studies, by exercise type | 18/70 (26%) |
| Queries with the stage relaxed (notice shown) | 29/350 (8.3%) |
| Typed entries mapped to the correct option, held-out set | 54/57 (94.7%) |
| Unrelated typed entries mapped to an option, held-out set | 1/10 (10%) |
| Query time, median (95th percentile) | 0.02 s (0.08 s) |
| Summary time, median (95th percentile) | 0.76 s (1.24 s) |
| *Reliability* |  |
| Queries meeting all eight consistency rules | 440/440 (100%) |
| Comorbidity coverage relative to the best achievable (queries with an eligible study) | 1.00 (154 queries) |
| Queries returning every exact-match study that fitted | 365/365 (100%) |
| Blood pressure entries handled correctly (stage or none) | 14/14 (100%) |
| Same studies on rerun | 40/40 (100%) |
| Same studies with input variants | 40/40 (100%) |
| Agreement of the blood pressure filter with the quality control rule (κ) | 0.77 (95.6% of names) |
| *Faithfulness (30 summaries)* |  |
| Summaries with no ungrounded number | 30/30 (100%) |
| Significance statements contradicting a p-value | 0/27 |
| Summaries referring to a protocol that was not selected | 0/30 |
| Summaries with the required AI disclosure | 30/30 (100%) |
| Summaries without prescriptive wording | 29/30 (96.7%) |
| Summaries within the length limits | 30/30 (100%) |
| Summaries without headings or tables | 30/30 (100%) |
| Summaries naming an exercise type not among the selected protocols | 4/30 (13%) |

Test queries: 440 (every stage and an unknown stage with each comorbidity option and each exercise category, plus 50 random combinations of two or three comorbidities), five results each. The held-out set of typed entries (67) was written and frozen before the synonym lists and the similarity cutoff were set. The 30 summaries cover selections of one, two and three protocols (ten each).
AI, artificial intelligence; κ, Cohen’s kappa.

Comorbidity queries always returned five studies (370 of 370). The portal first lists the studies that report a requested comorbidity and then fills the remaining places with the studies whose comorbidity profiles are most similar to the request. To check that it lists the right studies, we compared it with a separately written program that finds, directly from the PAHD table, the five studies of the requested stage that together report the most requested comorbidities. For all 154 queries for which at least one study of the stage reported a requested comorbidity, the portal’s list covered the requested comorbidities as well as this best possible list, and it included every study with an exact match that fitted in the list. For the other 211 comorbidity queries (58%), no study of the stage reported the comorbidity, and all five places were filled with similar studies.

Exercise-type queries returned five studies in 18 of 70 cases (26%). This reflects the database rather than a fault: the portal does not fill the list with other types of exercise, and for the less common types fewer than five studies exist at a given stage. When no study of the requested stage used the requested exercise type, which happened in 29 of the 350 queries that specified a stage (8.3%, all of them exercise-type queries), the portal searched the studies of all stages instead and informed the user.

Users can also type a comorbidity or an exercise type in their own words instead of choosing it from a list, for example “T2DM” or “heart attack”. The portal maps such an entry to the closest option, or ignores it if no option is close enough. We tested this mapping on 67 typed entries that were written in advance and kept apart while the mapping was adjusted: 57 entries that have a correct option and 10 unrelated entries, such as “broken arm” or “watching TV”. The portal mapped 54 of the 57 entries (94.7%) to the correct option and ignored 9 of the 10 unrelated ones.

The remaining checks also passed. All 14 test BP readings received the right stage, including values at each cut-off, and the two readings that cannot be read received none. Every query met all eight consistency rules, such as matching the requested stage and exercise type, listing no study twice and showing the same values as the PAHD table. The same studies were returned when a query was repeated, typed in a different case or spacing, or given with its labels in a different order (40 of 40). The portal’s BP filter agreed with the definition of resting SBP and DBP used in quality control for 95.6% of outcome names (Cohen’s κ = 0.77). The filter is deliberately broader, as it also includes ambulatory, home and central readings.

The AI summary is written by an LLM (Qwen3.8[32]) from a fixed prompt that contains only the data of the protocols that the user selected; the LLM explains the evidence in general terms but does not select or rank studies (Methods). Because such text could misstate the evidence, we generated 30 summaries for selections of one, two and three protocols (ten each), balanced between significant and non-significant results, and checked each of them by rule. First, all 99 numbers in the summaries appeared in the protocol data given to the model. Second, none of the 27 statements about whether a result was significant contradicted its p-value. Third, no summary referred to a protocol that had not been selected, but four summaries (13%) named an exercise type, aerobic or high-intensity interval training, that was not among the selected protocols, as general background, which the prompt allows. Fourth, all summaries ended with the required disclosure that the text is AI-generated, stayed within the length limits and had no headings or tables. One summary (3.3%) used prescriptive wording (“when prescribing exercise”), which the prompt forbids. These tests show that the portal behaves as specified. They do not show that its output improves clinical decisions.

## Discussion

To our knowledge, PAHD is the first structured database of PA interventions for hypertension. We built it with an LLM-assisted pipeline and paired it with a clinician-facing portal for profile-specific evidence retrieval. The pipeline combines two-step screening with a six-step, evidence-grounded extraction procedure, and it converts heterogeneous papers into a uniform record: one row per reported comparison. The resulting resource spans the demographic, comorbidity, medication, and FITT dimensions relevant to individualized prescription, and its extracted effects recover the expected antihypertensive signal of exercise, supporting face validity.

Three design choices drive the quality of the resource. First, decomposing extraction by the parts of a study (its structure, interventions, arms and results) keeps each model call scoped to a small, unambiguous slice of the paper, which we found necessary to obtain reliable per-arm and per-outcome values. This design also yields stable identifiers that make assembly deterministic and auditable. Fixing the structure of each study first, with two LLMs cross-checking each other and a third LLM adjudicating the disagreements that remained, limits structural errors in the later steps. Second, pairing every extracted value with a verbatim source quote grounds the output in the text and provides a direct handle for verification, which is important because modern LLMs cannot be assumed to be free of hallucination[33]. Third, standardization and quality control are deterministic and logged, and clear errors such as reversed signs were corrected by rule or by review against the paper. Against the extraction benchmarks, the pipeline recovered the structure of the studies almost perfectly and BP results with an F1 score of 90.3%, while the weaker fields point to concrete targets for refinement.

Compared with prior manually curated resources such as PAHFKB, PAHD reduces the human effort required to screen and extract from a large literature and is designed to be re-run as new trials appear, addressing the timeliness limitation of static, hand-built knowledge bases. Compared with LLM pipelines built for a single review question, PAHD keeps the arm-level protocols, baseline characteristics and results of every included study in one reusable table. Relative to general-purpose LLM assistants, the portal grounds its output in retrieved, peer-reviewed evidence and restricts the language model to explaining rather than selecting studies, which improves traceability at the cost of the fluency an unconstrained model might offer.

Our study has several limitations. First, the corpus is restricted to English-language, randomized controlled trials indexed in PubMed and published from 2005 onward, so relevant evidence in other designs, languages, or sources is not represented, and studies without accessible full text were excluded. Second, only the study structure was cross-checked by two LLMs; the other steps rely on a single LLM. Third, the benchmarks are modest in size (36 test studies for extraction), each relied on a single adjudicator, and results were evaluated for BP outcomes only, so the accuracy for other outcomes is unknown. Fourth, several fields are extracted less accurately and should be interpreted with caution: the number completing each arm, exercise intensity, and the dispersion and number analyzed of BP results. Fifth, the portal’s matching is rule-based and uses only the hypertension stage, comorbidities and exercise category. Its summaries cover the protocols that the user selects, may add general background that is not in the data, and in one of 30 tests used prescriptive wording. As this study primarily focused on developing the evidence database and establishing the technical feasibility and reliability of the portal, its evaluation was limited to automated testing. Formal clinician evaluation and assessment of patient outcomes were beyond the scope of the present study and remain important directions for future research. Finally, PAHD is a research and clinical-decision-support resource, not a medical device; its output summarizes existing evidence and is not a substitute for individualized clinical judgment.

Several directions follow naturally. The corpus could be broadened beyond English-language randomized controlled trials and refreshed on a schedule to keep the database current. Importantly, the database provides an evidence-grounded foundation for developing an interactive patient–clinician chatbot that integrates clinical characteristics with patients’ exercise preferences, motivations, and physical limitations to support clinicians in delivering reliable, personalized physical activity recommendations. More broadly, the pipeline is not specific to hypertension: because it is organized around the generic structure of a randomized controlled trial, the same framework could be applied to other chronic conditions in which physical activity is therapeutic, such as obesity, type 2 diabetes, and heart failure, laying the groundwork for scalable, personalized, evidence-based guidance across chronic disease management.

## Methods

### Data sources and corpus construction

PubMed was selected as the primary data source given its broad coverage of peer-reviewed biomedical literature and high-quality metadata. To scope the database, we first specified PICO terms[34]: the population is people with hypertension; the intervention involves physical exercise; the comparison is a no-intervention or sedentary control versus exercise; and the outcome includes, at minimum, blood pressure. Expanding these terms with synonyms and related concepts produced a Boolean query that combines three groups of terms (hypertension; exercise or control conditions; blood pressure) with AND, and the terms within each group with OR (Table A in S1 Appendix). We then applied filters on publication date, text availability, article type, language, species, and participant age. We retained studies published between 2005/01/01 and 2025/12/31 with accessible abstracts and full texts, restricted to randomized controlled trials, written in English, and conducted in adults (≥19 years, per PubMed age-group definitions). Randomized controlled trials were chosen because they provide well-defined exercise protocols and study populations; meta-analyses and systematic reviews were excluded because they summarize rather than report the arm-level granularity the database requires. The resulting study metadata (PMID, title, abstract, MeSH terms, journal, publication year, and publication type) formed the input to screening. Exact record counts at each stage are reported in Results and Fig A in S1 Appendix.

### Study screening

Screening used an LLM (OpenAI gpt-6-luna) in two steps with the same five eligibility criteria (prompts in Figs B–D in S1 Appendix). We chose gpt-6-luna, for screening and for most extraction steps, to balance capability and cost, given the large number of calls. The criteria asked whether: (1) the study population included individuals with hypertension or elevated BP (prehypertension); (2) a structured exercise training program with multiple sessions over days or weeks was evaluated as an intervention; (3) the effect of exercise could be isolated from other intervention components, such as diet, for the hypertensive population; (4) the study was a completed intervention study with reported results; and (5) BP (e.g., SBP, DBP, MAP, ambulatory BP) was reported as an outcome, with a statistically supported comparison involving the exercise arm. For each criterion the model returned a decision and a supporting quote, plus a one-sentence explanation for any decision other than “Yes.”

The title-and-abstract step used only the title and abstract, with the model’s extended reasoning turned off (reasoning effort “none”). The model could answer “Yes,” “No,” or “Unclear.” It was told to prefer “Unclear” whenever the abstract was silent or ambiguous, because a wrong “No” removes a study for good. Studies with at least one “No” were excluded. The full-text step used the full text of the paper, with the title-and-abstract decisions as context only, and a low reasoning effort. Because the API cannot read PDF files directly, each paper was first converted to Markdown with the Mathpix API[35]. Mathpix was chosen because it reliably converts tables and mathematical expressions with sufficiently high quality; most results in our corpus are reported in tables, and a poor conversion of tables would cause values to be lost or attributed to the wrong group. The model re-judged every criterion and had to answer “Yes” or “No.” A study was retained only if all five criteria were “Yes.” Both steps followed the same decision rules. For example, a study of the acute response to a single exercise session failed criterion 2. A multi-arm trial passed criterion 3 if one arm differed from another only by exercise. Statistical support for criterion 5 could be a p-value, a confidence interval, a test statistic, a significance symbol defined in a table legend or footnote, or an explicit statement of significance or non-significance, provided that the paper states its significance level.

### Entity-relationship data model

To handle heterogeneity in how studies present their results, we defined the target fields with an entity-relationship model (Fig 2). A Paper holds study-level facts: design, setting, eligibility criteria, sample size and the significance threshold. An Arm is a group of participants that the paper allocates or recruits as a distinct group. It holds the group’s size and its baseline characteristics: age, sex, race and ethnicity, BMI, comorbidities, medications and baseline BP. An Intervention is a condition that participants receive, such as an exercise program or a control condition. It holds the verbatim protocol and its prescription parameters: the FITT components of frequency, intensity, time (session duration) and type (modality), plus total program duration, supervision and progression. Links record which arm receives which intervention. A Time point is a moment of measurement, with baseline as the reference. An Outcome is a measure assessed to evaluate the intervention. A Result is one comparison between two sides, where a side is a set of arms and interventions at a time point. It holds the statistic of each side, the difference between the sides and the p-value. A comparison can follow one arm over time or compare groups. Each Result is one record of the database.

### Information extraction

Extraction ran in six steps: *skeleton*, *outcomes*, *study*, *interventions*, *arms* and *results*. Each step had its own prompt (Figs E–O in S1 Appendix) and a fixed JSON schema. Together, the steps collect the data items that the Cochrane Handbook lists for data collection in systematic reviews[23]: methods, participants, interventions, outcomes and results. We did not ask a model to extract a whole paper in one pass. Research papers are hierarchical: a study defines arms, interventions, time points and outcomes, and every result is indexed by them. Enumerating all of these while also filling in every value imposes a large combinatorial load, which in our experience degrades accuracy and increases misattributed values. We therefore fixed the structure of each study first (the skeleton and outcomes steps). Later steps received this structure as compact entity cards with stable identifiers, and each call was scoped to one paper, one intervention or one outcome. The identifiers also make the later joins deterministic and auditable.

Every request contained the whole paper: the Markdown text produced by Mathpix (Study screening) and, as images, the figures of the paper, which Mathpix extracts separately. Each figure was attached after the text, and a marker at its original position in the text (for example, “attached_image_2”) told the model where it belonged. The model could therefore read values that are shown only in a figure, such as BP over time or the numbers in a participant flow diagram. Such values carry the figure as their location, and their quotes cannot be checked against the text.

The skeleton step identifies the study skeleton: the design, the arms, the interventions, and which arm receives which intervention. Arms must be disjoint and must together cover all allocated participants; pooled groups and subgroups are not arms. For each arm, the model also lists every string under which the arm appears in the text and tables, so that later steps can locate its table columns. The outcomes step identifies the outcomes and the time points at which each outcome is reported. An outcome counts only if the paper reports results for it in its main text, tables or figures. Measurement variants, such as office, 24-hour, daytime and central SBP, are kept as distinct outcomes. Baseline-only descriptors, adverse events and process measures such as adherence are excluded.

Errors in the skeleton or the outcome list propagate to every later step. The skeleton and outcomes steps were therefore run with two independent extractors, OpenAI gpt-6-luna and Google gemini-3.8-flash, both with a medium reasoning effort. A program then aligned the two outputs. Arms were matched one to one by the similarity of their names (including every name under which an arm appears in the paper), equal numbers of participants, the similarity of the interventions they receive and their position; interventions by their names and by the arms that receive them; and outcomes and time points mainly by their names and the table labels under which they appear, never matching, for example, a systolic with a diastolic outcome. Each disagreement between the aligned outputs became an item, for example a different design, an arm or outcome that only one model lists, a different division of the study into arms, an uncertain match, a different number of participants in an arm, a link between an arm and an intervention that only one model lists, or a time point that only one model treats as the baseline. In a cross-check, both models reviewed every item blindly: they saw the two answers in random order, without knowing which model had written which answer, and judged them against the paper. An item was resolved when the two reviewers agreed. Otherwise, it went to a final adjudicator (Anthropic Claude Sonnet 5.5), which weighed the answers and the reviewers’ arguments against the paper text. Disagreements were uncommon. In the skeleton step, 23 of 255 papers had at least one disagreement (47 items; 20 resolved by the cross-check and 27 by adjudication). In the outcomes step, 115 papers had at least one disagreement (357 items; 256 resolved by the cross-check and 101 by adjudication). The final skeleton and outcome list of each paper were then assembled by program.

The four other steps extract the details against this structure with gpt-6-luna (medium reasoning effort). The study step extracts facts about the study as a whole: design, countries, number of centers, numbers enrolled and randomized, eligibility criteria, the definition of hypertension, statistical methods and the significance threshold. The interventions step runs once per intervention. It copies the complete protocol description verbatim, classifies the intervention type (e.g., exercise, usual care or no intervention) and whether the paper frames it as a control, and extracts its frequency, session duration, intensity, modality, total duration, progression, supervision and equipment. The arms step extracts, for each arm, the number completing the study and the baseline characteristics (age, sex, race and ethnicity, BMI, comorbidities, medications and baseline BP) as reported, with their statistic type and dispersion. The results step runs once per outcome. It extracts every comparison reported for that outcome, across all arms, interventions and time points. For each side of a comparison, it records the statistic, its dispersion and the number analyzed. For the comparison, it records the reported difference and the p-value. The model also receives the table footnotes, so that significance symbols can be resolved.

All steps follow the same grounding rules. Every extracted value is paired with a verbatim evidence quote and its location in the paper. The model must keep the paper’s own wording, numbers and units, and must not convert, compute or infer values; normalization is left to the deterministic steps that follow. A field can carry a status (“not reported,” “not applicable” or “unclear”) instead of a value, so that missing information is not guessed. A program checks each quote against the paper text, allowing for formatting differences such as LaTeX markup. The output of the results step is also checked against the entity cards and the paper: every arm, intervention and time point that a result cites must exist and be linked to the outcome, each arm must receive the interventions listed with it, a result must not compare a side with itself, every number must have a supporting quote that is found in the paper, and the same group must not carry different values in different rows. Failed checks are logged as warnings rather than fixed; the quality control step later turns some of them into flags, for example a time point that is not linked to the outcome.

### Database construction

The database construction stage turned the extraction outputs into the database in four deterministic steps, without further LLM calls. The tables step flattened the outputs into linked tables, one per entity: papers, arms, interventions, time points, outcomes, baseline statistics and results. The tables are keyed by the stable identifiers and keep the raw values and their evidence.

The standardization step filtered and standardized these tables. It removed result rows that compared two baseline values or held no result, comparisons between two control conditions or between medication-only arms, the one study in which no arm received exercise, and one study that reported BP only as correlations with other measures. Each intervention was mapped to a PA category: aerobic, resistance, flexibility, neuromotor (e.g., yoga or tai chi) or other forms (e.g., breathing exercise or inspiratory muscle training). Non-exercise conditions were labeled by type, such as usual care or no intervention. Frequency and session duration were expressed in consistent units, and a weekly exercise volume was computed from them. These PA mappings were made by rules and then reviewed by hand for every intervention. Countries were mapped to ISO 3166-1 alpha-3 codes[36]. Baseline characteristics were mapped to standard vocabularies: race and ethnicity to the US OMB categories[37], medications to WHO ATC classes[38] and comorbidities to ICD-10 titles[39]. P-values, however written (e.g., “P<0.001,” “NS”), were parsed into an operator and a number; when a paper stated no significance threshold, 0.05 was assumed.

The joining step joined the tables into one flat table with one row per reported comparison. Each row carries the study context, the protocol and baseline characteristics of each side, the statistics of each side and the contrast. Rows were oriented so that the exercise side comes first, and differences follow one sign convention: intervention minus comparator, and later minus earlier time point. Each arm was assigned BMI categories (WHO cut-offs[40]) and a hypertension stage (2025 AHA/ACC guideline[3]) from its mean baseline values. Comparisons reported more than once, for example in a table and in the text, were merged when compatible and otherwise kept as distinct results; the few conflicting duplicates were resolved by hand.

The quality control step checked the joined table record by record. Rules tested each record for internal consistency, and each failed test adds a named flag: the reported difference does not match the two group values; the direction of a difference cannot be determined; an interval does not contain its estimate, or contradicts the p-value; a number does not appear in the evidence quotes or the paper; a resting BP value is implausible; the number analyzed exceeds the number assigned; a value repeated in several places differs; the stated direction contradicts the values; or a standard deviation, standard error or interquartile range is negative. Clear errors with a deterministic fix, such as a reversed sign convention or swapped interval bounds, were corrected automatically, and the value as reported was kept in the record. Flags on BP outcomes were reviewed by hand against the paper. The reviewer could use a general-purpose LLM chat assistant (for example, ChatGPT) to locate information in a paper or to explain symbols and footnotes, but made every decision and was responsible for the result. In the final table, 15,389 records (89%) passed every check and 390 (2%) were corrected by rule, mostly to harmonize the sign convention of reported differences. Of the flagged BP records, 392 were confirmed or corrected by hand and 55 were removed. The remaining 1,102 records (6%), 89% of them on non-BP outcomes, keep an open flag. Numbers that an LLM read from a figure image cannot be checked against the text, so a separate column marks the 169 records in which some numbers come only from a figure. Every fix, edit and open flag is recorded in the quality-control status and flag columns of the database.

### Screening validation

We built a screening benchmark of 500 studies from the pool of 2,548. To include enough eligible studies, sampling was stratified by keyword flags in the title, abstract and MeSH terms. Stratum S1 held studies with both a hypertension and an exercise term (880 in the pool; 225 sampled), S2 an exercise term only (920; 200) and S3 no exercise term (748; 75). Each study carries a stratum weight, the inverse of its stratum’s sampling fraction (3.9, 4.6 and 10.0), so that weighted estimates describe the whole pool.

Two annotators judged each study independently on the same five criteria, blinded to the stratum and to the pipeline’s decisions. A human adjudicator resolved every disagreement against the paper. The annotators and the adjudicator could use general-purpose LLM chat assistants, for example to locate information in a paper or to explain symbols and footnotes, but made every judgment themselves and were responsible for the final labels. We summarized agreement before adjudication with raw agreement, Cohen’s κ and Gwet’s AC1.

We scored the end-to-end screening decision: a study counted as included only if it passed both screening steps. We report recall, precision, F1, F2, specificity and work saved, the proportion of the pool not passed on to extraction. All estimates are weighted by the stratum weights, with 95% confidence intervals from a stratified bootstrap of studies. We also report the title-and-abstract step alone, and results for studies whose label was agreed or adjudicated (Note C in S1 Appendix).

### Extraction validation

The extraction benchmark comprised 50 studies drawn from the included set, stratified by type of exercise so that every type is represented. One was a meta-analysis and was excluded, leaving 49. Thirteen studies had been used during prompt development (development set). The other 36 form the held-out test set, our primary analysis set. Two annotators annotated each study independently with the same workbook, field definitions and PDF. Their annotations were aligned record by record. A human adjudicator then resolved every disagreement cell by cell against the paper, choosing either value or typing a correction. As for the screening benchmark, the annotators and the adjudicator could use LLM chat assistants in this way, but every value and decision was theirs. Results were adjudicated for BP outcomes only (SBP, DBP, mean arterial pressure, pulse pressure and their variants), so the evaluation of the results step is restricted to them.

The pipeline names entities independently of the benchmark. Before any value was compared, each extracted arm, intervention, time point and outcome was therefore matched one-to-one to a benchmark entity, using names, linked entities, sample sizes and position. The adjudicator reviewed every uncertain or unmatched pair among the arms, interventions, time points and BP outcomes. Values were compared after normalization, using documented rules that equate only formatting differences (e.g., “p<0.01” and “P < .01”).

For specific extracted entities, we report precision, recall and F1. For study- and arm-level fields, we report end-to-end accuracy, in which a value of a missed entity counts as wrong, and macro-F1 for closed lists with rare classes. The FITT parameters are free text. A pair counted as correct when the texts were equal after normalization, the extraction contained the benchmark text, or their numbers and units matched. The remaining pairs were judged by an LLM (gemini-3.8-flash). This judge was not validated against a human rater, so we also report bounds that count every judged pair as wrong or as correct. To check where the verbatim protocol passages came from, we located each extracted and each benchmark passage in the Markdown text of the paper, by covering it with matching stretches of at least 20 characters after normalization, and assigned each located stretch to its paragraph and, through the Markdown headers, to its section. We report the proportion of each passage that was found, the sections it came from, and the overlap between extracted and benchmark passages by paragraph and by character (Note D in S1 Appendix). BP results were scored as data points rather than table rows: the statistic of one group at one time point (or its change), and each between-group contrast. A data point was correct when its outcome, groups and time points matched and its value was equal. We also report the proportion of benchmark data points whose value and dispersion were both correct (meta-analysis-ready), and the proportion of extracted values printed in the paper (grounding). Each benchmark data point that was not meta-analysis-ready was assigned one error category: a dispersion error (the value is correct, but its dispersion is missing or different), a sign error (the benchmark value with the opposite sign), a misattribution (the value was extracted, but for another group, time point or outcome), a transcription error (the data point was extracted with a different value) or an omission (the value was not extracted at all).

Confidence intervals are 95% percentile intervals from a paper-level cluster bootstrap with 2,000 replicates. In a sensitivity analysis, we removed the entities that the adjudicator judged to be defensible alternative readings rather than errors. In an ablation, we scored each single extractor (gpt-6-luna, gemini-3.8-flash) of the skeleton and outcomes steps and compared it with the cross-checked output. Both are reported in Note D in S1 Appendix.

### Web portal

The web portal is a Shiny for Python application[41] that searches the PAHD table. Its search pool contains the comparisons that involve an exercise arm and a blood pressure level measured at rest, over 24 hours (ambulatory), at home or centrally (3,117 comparisons from 248 studies). Blood pressure outcomes that are not such a level, such as responses during an exercise test, changes reported as separate outcomes, variability and the proportion of patients reaching blood pressure control, were left out by matching their names against a list of words. For each study the portal shows one result, chosen in this order: the kind of blood pressure outcome (standardized SBP or DBP, then other resting or office blood pressure including mean arterial pressure, then ambulatory, home or central blood pressure); the kind of comparison (exercise against control, then the exercise arm over time, then exercise against another exercise arm); a single arm before a pooled set of arms; systolic before diastolic before mean pressure; the primary analysis before an intention-to-treat analysis; the full sample before a subgroup; and finally the significance of the p-value (p < 0.05 first, then the smaller p-value). A study’s hypertension stages are those of its arms (2025 AHA/ACC stage of the mean baseline blood pressure), and its comorbidities are the ICD-10 labels in PAHD. The journal’s Journal Impact Factor is taken from Clarivate’s Journal Citation Reports and is shown as not available for journals without one.

The user sets the number of studies to show (one to five), the patient’s stage and the matching mode. The stage is selected, or derived from a typed blood pressure reading with the 2025 AHA/ACC cut-offs[3] (Stage 2: systolic ≥140 or diastolic ≥90 mm Hg; Stage 1: 130–139 or 80–89; elevated: systolic 120–129 and diastolic <80; otherwise normal). Matching is rule-based. The pool is first restricted to studies that include the stage; if there is none, all stages are searched and the user is informed. In comorbidity mode, studies that report at least one selected comorbidity come first, ordered by the number of selected comorbidities they report and then by significance. If there are fewer studies than requested match, the list is completed with the remaining studies whose comorbidity profile is most similar to the query, measured by the cosine similarity of sentence embeddings (all-mpnet-base-v2)[42,43]. In exercise-type mode, studies with a selected exercise category (aerobic, resistance, flexibility, neuromotor or other) are ordered by significance. Age, sex, race, region, body mass index and medication are not used for matching.

Users can type comorbidities and exercise types instead of choosing from the lists. A typed entry is embedded and compared with the embeddings of each option label and of a short list of synonyms, abbreviations and lay terms written for it (for example “T2DM” or “heart attack”). The best option is used if the cosine similarity is at least 0.55. Otherwise, the closest option by fuzzy string matching (similarity of at least 0.6) is used, and an entry without a match is ignored. The synonym lists and the 0.55 cutoff were written after a first set of 58 typed entries showed mapping errors; the mapping was then tested on a second set of 67 entries that had been written, and frozen, before these changes.

The user can select some of the displayed protocols and request a summary. A fixed prompt gives the language model, for each selected protocol, the title, journal and impact factor, the exercise category, frequency, intensity, session duration and modality, the blood pressure outcome and its comparison, the p-value with its stated significance, the change in the statistics and the type of comorbidity match. The model (Qwen3.8, qwen/qwen3.8-27b[32], served by Groq[44]; temperature 0.1; at most 400 tokens) is asked for 120 to 170 words that explain in general terms why this type of exercise lowers blood pressure, what the statistics mean, the trade-offs between protocols and a brief safety note. It must not repeat the protocol specifications, invent protocol-specific facts or reinterpret p-values, must call the items “protocols” rather than recommendations, and must end with a fixed sentence that discloses that the text is AI-generated. The language model has no role in selecting or ranking studies.

### Web portal evaluation

We evaluated the portal with code, without human raters or an LLM judge. The test code calls the functions that the server uses to run a query and to build and request a summary. The 440 test queries (five results each) combined each of the four stages and an unknown stage with each of the 64 comorbidity options (including none) and each of the 14 exercise categories, plus 50 random combinations of two or three comorbidities with a random stage.

For usability, we recorded the proportion of queries that returned the requested number of studies and the proportion for which the stage had to be relaxed. Typed-entry mapping was scored on the second set of 67 entries (57 with a correct option and 10 without) as top-1 accuracy, the proportion of entries with a correct option that were mapped to it, and as the false-mapping rate, the proportion of entries without a correct option that were mapped to an option anyway. Query and summary times were compared with response-time limits of 1 s and 10 s.

For reliability, every query was checked against eight rules: the portal returns at most the requested number of studies; it lists no study twice; it honors the requested stage, unless no study of that stage exists, in which case it says so; it returns only studies of the requested exercise type; it lists studies that report a requested comorbidity before similar ones; it orders the studies as documented; every row has a blood pressure outcome; and every displayed value equals the value in the PAHD table. We also compared the coverage of the requested comorbidities with that of an independent program that works directly on the PAHD table. The coverage of a study is the proportion of the requested comorbidities that it reports, for example 0.5 for a study that reports one of two requested comorbidities. For each query, attainment is the summed coverage of the returned studies divided by the largest summed coverage that any set of studies of the stage could reach; an attainment of 1 means that no better list exists. We also checked that every study with an exact comorbidity match was returned whenever there was room for it. Stage assignment was tested on 14 typed blood pressure readings: values just below, at and above each cut-off, readings in different formats, and two readings that cannot be read.

The portal’s blood pressure filter was compared, outcome name by outcome name, with the definition of resting SBP and DBP used by the quality control step (Cohen’s κ). Determinism was tested on 40 sampled queries: each was repeated, typed with its labels in a different case or spacing, and given with its labels in a different order.

For faithfulness, we generated 30 summaries for selections of one, two or three protocols (ten each) from the test queries, balanced between significant and non-significant results, and checked them by rule. We extracted every number from each summary and checked that it appears in the protocol data given to the model; a rounded form of a number in the data (for example, 10.6 for 10.62) counted as a match. A statement of significance attributed to a protocol had to agree with its p-value, every protocol or PMID mentioned had to be one of the selected ones, and every exercise type mentioned had to appear in a selected protocol. A summary had to end with the disclosure sentence, contain no prescriptive wording (such as “recommend,” “prescribe” or “you should”), have 120 to 170 words (with a tolerance of 20%) and contain no headings or tables.

The evaluation tests whether the portal behaves as specified, not whether its output improves clinical decisions.

## Data Availability

The data will be available online at Zenodo

## Ethics and intended use

This study used only data reported in published articles and involved no human participants; ethical approval and informed consent were therefore not required. PAHD is a research and clinical-decision-support resource intended to summarize published evidence for clinicians; it is not a medical device and does not provide individualized medical advice. Its output should be interpreted alongside, and not as a substitute for, clinical judgment.

## Supporting information

**S1 Appendix. Supplementary notes, figures and tables.** Notes A–F, Figs A–O (PRISMA flow diagram and LLM prompts) and Tables A–B (PubMed search query; LLM calls, cost and runtime per step).

