## Supplementary material for "A knowledge database of physical activity interventions for hypertension using large language models": Note A-F, Fig A-O, Table A-B

**S1 Appendix. Supplementary notes, figures and tables**

**Contents**

**Notes**

**Note A.** Study selection

**Note B.** Information extraction run

**Note C.** Screening accuracy: workload and errors

**Note D.** Extraction accuracy: further analyses

**Note E.** Contents of PAHD at the study level

**Note F.** Studies in the search pool of the web portal

**Figures**

**Fig A.** PRISMA flow diagram of the selection of studies for PAHD

**Fig B.** System prompt for screening

**Fig C.** Prompt for title-and-abstract screening

**Fig D.** Prompt for full-text screening

**Fig E.** System prompt for information extraction

**Fig F.** Prompt for the study skeleton

**Fig G.** Prompt for cross-checking skeleton discrepancies

**Fig H.** Prompt for adjudicating skeleton discrepancies

**Fig I.** Prompt for outcomes and time points

**Fig J.** Prompt for cross-checking outcome and time point discrepancies

**Fig K.** Prompt for adjudicating outcome and time point discrepancies

**Fig L.** Prompt for study-level facts

**Fig M.** Prompt for intervention details

**Fig N.** Prompt for arm details and baseline characteristics

**Fig O.** Prompt for the results of one outcome

**Tables**

**Table A.** PubMed search query and filters

**Table B.** LLM calls, cost and runtime per step

**Note A. Study selection**

The 461 studies that passed the title-and-abstract step had either only “Yes” answers (189) or a mix of “Yes” and “Unclear” (272). In the full-text step, the LLM excluded 198 of the 453 studies whose full text could be retrieved. Counting each excluded study once, under the first reason that applied in the order listed here, 4 were not completed intervention studies, 102 did not study a hypertensive or elevated-BP population, 23 did not evaluate a structured, multi-session exercise program, 43 did not allow the exercise effect to be isolated, and 26 did not report a statistically supported BP comparison involving an exercise arm (Fig A).

**Note B. Information extraction run**

Of the 255 papers, 239 had at least one figure attached (705 figures in total). Across all steps, the pipeline produced 53,782 evidence quotes. Code found 98.9% of the 52,187 quotes taken from the text in the paper itself: 93.0% verbatim and 5.9% after ignoring formatting differences such as LaTeX markup. The other 1,595 quotes, in 198 papers, were read from figure images; for example, 1,059 of the 19,476 extracted results, in 107 papers, used values read from a figure.

**Note C. Screening accuracy: workload and errors**

In the screening benchmark, the title-and-abstract step alone kept 91.7% of eligible studies while passing on 18% of the pool to the full-text step. The 251 records that the pipeline passed on in total correspond to 1.24 studies for every eligible study found.

The pipeline missed 8 of the 59 eligible benchmark studies and wrongly kept 11 ineligible ones. Five eligible studies were dropped at the title-and-abstract step. In four of them, the abstract did not report a statistically supported BP comparison, and the LLM answered “No” instead of “Unclear” on criterion 5 (BP reported as an outcome, with a statistically supported comparison involving an exercise arm); the fifth evaluated device-guided breathing, which the LLM did not count as exercise training. Three were dropped at the full-text step because the LLM judged that their BP comparisons, which were not statistically significant, did not meet criterion 5, although a stated non-significant result meets it. All 11 wrongly kept studies failed the population criterion in the benchmark: most enrolled broader groups, such as people with type 2 diabetes, metabolic syndrome or obesity, of whom only some had hypertension, and did not analyze them as a separate group. Errors concentrated on studies that were hard for the annotators as well: 17 of the 19 errors concerned studies on which the two annotators had disagreed and the adjudicator decided. Among the studies on which both annotators agreed, recall was 100% and precision 92.0%.

**Note D. Extraction accuracy: further analyses**

**Cross-checking of the study structure.** To see what cross-checking added, we also scored the structure that each LLM extracted on its own, before the cross-check, and compared it with the cross-checked structure; for this comparison, the outcomes of each LLM alone and of the cross-checked list were all matched to the benchmark by their names only, which is why the cross-checked outcome F1 below (92.3%) differs from the 94.0% of the main analysis (Table 3). Cross-checking mainly helped the arms and the BP outcomes. The F1 for BP outcomes rose from 97.7% (gpt-6-luna alone) and 98.6% (gemini-3.8-flash alone) to 99.4%, and the proportion of studies with exactly the right set of arms rose from 97.2% (gpt-6-luna alone) to 100%. For all outcomes, the cross-checked list (F1 92.3%) was better than that of gpt-6-luna alone (90.7%) but not better than that of gemini-3.8-flash alone (94.8%).

**Study facts and baseline statistics.** The type of each intervention, one of ten categories such as exercise, exercise combined with another component, usual care or no intervention, was correct for 87.8% of interventions (Table 3), with a macro-F1 of 77.8%; most errors confused neighboring categories, such as exercise alone and exercise combined with another component. When a baseline statistic was found, its type of dispersion and the dispersion value were almost always correct (99.6% and 99.4%).

**Exercise prescription.** Whether each of the five prescription parameters was reported agreed well with the benchmark (Cohen’s κ = 0.83). Total program duration was the least accurate parameter (72.0%; Fig 3b), mostly because of a difference in convention: for 17 control arms that received no intervention, the benchmark recorded the length of the study period (for example, “12 weeks”) as the duration, whereas the pipeline recorded it as not applicable.

**Protocol passages.** The verbatim protocol passages came from the right part of the paper. We located every extracted passage, and every benchmark passage, in the text of the paper. All 81 extracted passages of the test set were found in the paper, and 99.8% of their text came from the Methods section. The extracted and benchmark passages came from the same paragraphs: 85.3% of the paragraphs that held a benchmark passage also held the extracted passage, and 87.3% of the paragraphs that held an extracted passage also held the benchmark passage. The extracted passages covered 94.0% of the text of the benchmark passages, but they were about twice as long, as only 52.9% of their text was also part of the benchmark passage; the rest also came from the Methods section.

**Blood pressure results.** Differences between groups were harder than group statistics (recall 71.9% of 32 data points). Of the 107 dispersion errors (Fig 4), in 59 the pipeline did not record the type of dispersion, typically because the paper stated it only once, in its statistical methods (for example, “values are expressed as mean and standard deviation”), rather than next to the numbers; in 22, the type was wrong, such as a CI recorded as a standard deviation; in 25, the dispersion value itself differed; and in one, the pipeline added a dispersion that the benchmark does not have. The number of participants analyzed in the group was correct for only 53.9% of the matched data points: the pipeline left it empty for 365 of the 797 matched data points for which the benchmark records it. Of the matched data points with a p-value in the benchmark or the extraction, the p-value was correct for 88.5% (300 of 339); of those with a statement of significance, the statement was correct for 83.8% (67 of 80).

**Sensitivity analysis.** Some differences between the pipeline and the benchmark were not errors but defensible alternative readings of a paper. In one study (PMID: 21985749), for example, the pipeline did not separate four weekly time points that were shown only in a figure, and in another study (PMID: 34347008), it kept nighttime ambulatory SBP and DBP as one outcome where the benchmark kept two. The adjudicator marked eight such entities, six of them in the test set. Leaving them out changed the results little: the F1 for BP data points in the test set was 89.9% instead of 90.3%.

**Note E. Contents of PAHD at the study level**

Most studies were parallel-group randomized trials (228; 90%), with 10 crossover, 4 factorial and 4 cluster-randomized trials. Studies contributed a median of 57 records (interquartile range [IQR] 37–86; range 4–352), reflecting wide variation in the number of arms, time points and outcomes per study. The median study randomized 48 participants (IQR 30–79; range 9–2,157).

The median of the study mean ages was 58 years (IQR 51–65; range 21–82). Sixty-five studies (26%) had a mean age of 65 years or more, and 25 (10%) a mean age below 45 years. Sex was reported in 97% of studies; 42 enrolled women only and 24 men only. Body mass index was reported in 90% of studies, and arms were most often overweight (72% of studies) or obese (56%) by WHO categories (Methods). Baseline BP allowed a 2025 AHA/ACC stage to be assigned in 62% of studies: Stage 1 hypertension in 33%, Stage 2 in 31% and elevated BP in 12%. Studies came from 41 countries, most often Brazil (12%), the United States (7%), China (4%), Nigeria (4%) and Germany (3%); 35% did not state a country. Race and ethnicity were reported in 22% of studies; where reported, the most common groups were White (13% of all studies) and Black or African American (9%).

Medication use was reported in 51% of studies. The leading classes were calcium channel blockers (25% of studies), ACE inhibitors (23%), diuretics (22%), angiotensin II receptor blockers (22%) and beta blockers (21%); 23% of studies also recorded antihypertensive use without a class. Comorbidities were reported in 42% of studies. Diabetes was the most common (25%), followed by metabolic syndrome (7%), dyslipidemia (7%) and obesity (6%).

Of the 368 exercise interventions, 15% combined aerobic and resistance training. Where a weekly volume could be computed (284 interventions), the median was 129 minutes per week (IQR 90–180). Intensity was described for 90% of interventions, and 52% were fully supervised.

**Note F. Studies in the search pool of the web portal**

Of the 248 studies in the search pool of the portal, 84 include Stage 1 hypertension, 78 Stage 2, 31 elevated BP and 7 normal BP, and 77 report at least one of 63 comorbidities. The BP filter of the portal is deliberately broader than the definition of resting SBP and DBP used in quality control, as it also includes ambulatory, home and central readings, but it left out 35 outcome names that the quality control definition accepts.

**Identification and selection of studies for PAHD**

Records removed *before screening*:

Records without access to abstract and full text (n = 1290)

Non-English and non-human records (n = 5411)

Records excluded due to other reasons (n = 15319)

Records identified from Pubmed within the time range:

(n = 24568)

**Identification**

Records screened by title and abstract

(n = 2548)

Records excluded

(n = 2087)

Reports sought for retrieval

(n = 461)

Reports not retrieved

(n = 8)

**Screening**

Reports excluded:

Ineligible population (n = 102)

No structured exercise training program (n = 23)

Exercise effect cannot be isolated (n = 43)

Not a completed intervention study (n = 4)

No eligible blood pressure comparison (n = 26)

Reports assessed for eligibility

(n = 453)

Studies eligible for information extraction

(n = 255)

**Included**

**Fig A. PRISMA flow diagram of the selection of studies for PAHD**

Records were identified in PubMed within the publication period (2005–2025), screened by an LLM on title and abstract and then on full text (Methods), and the eligible studies passed to information extraction. Each excluded report is counted once, under one reason.

You are a screening assistant. You read scientific papers (a title and abstract, or the full paper text) describing physical activity or exercise interventions, and you judge whether each paper is eligible for the PAHD database, which studies the effect of structured exercise training on blood pressure in hypertensive populations.

Use only the text and structured inputs provided by the user. Do not use outside knowledge, and do not assume details that are not explicitly stated.

You evaluate each eligibility criterion defined in the task instructions independently, choosing from the decision values that those instructions and the output schema allow. Apply the criteria, decision definitions, and tie-breaking rules exactly as the task instructions specify.

General rules:

Decisions

1. Judge each criterion independently, based only on what the provided text explicitly supports. Do not infer eligibility from indirect clues, outside knowledge, or assumptions about what the study probably did.

2. Ground every decision in the provided text. When the text supports the decision, give the shortest sufficient verbatim evidence quote; when no relevant text exists, set the evidence field to null.

3. Evidence quotes must copy the provided text verbatim as closely as possible. Whitespace may be normalized, but words must match. Do not paraphrase or fabricate evidence.

4. Keep evidence quotes short. Do not copy entire tables or long blocks unless absolutely necessary.

5. When the text is insufficient to decide a criterion, follow the task instruction's guidance for that case rather than guessing.

6. For "No", "Unclear", or any decision made on an ambiguous reading, give a concise one-sentence explanation grounded in the text.

7. Return the requested structured JSON only, conforming to the provided schema. Do not add any text outside the JSON.

System prompt for screening

**Fig B. System prompt for screening**

System prompt of the screening stage. It was the system message of every title-and-abstract and full-text screening request (OpenAI gpt-6-luna), sent together with the task prompts of Figs C and D. Shown verbatim.

Task: Step 0a title/abstract screening.

This is the first pass of paper screening. Decide whether a paper potentially meets the eligibility criteria for a study examining the effect of structured physical exercise training on blood pressure in hypertensive populations, using only the paper title and abstract. The purpose is to exclude only papers that are clearly out of scope and to pass anything uncertain forward to full-text screening.

You will receive:

1. paper_id

2. Paper title

3. Paper abstract

Return a JSON object conforming to the provided schema.

Eligibility criteria:

1. Does the study population include individuals with hypertension or elevated blood pressure (prehypertension)?

2. Is a structured exercise training program (multiple sessions over days or weeks) evaluated as an intervention?

3. Can the effect of exercise be isolated from other intervention components (e.g., diet) for the hypertensive population?

4. Is this a completed intervention study with reported results?

5. Is blood pressure (e.g., SBP, DBP, MAP, ambulatory BP) reported as an outcome, with a statistically supported comparison involving the exercise arm?

Decision values:

- "Yes": the title or abstract clearly satisfies the criterion.

- "No": the title or abstract clearly fails the criterion.

- "Unclear": the title and abstract do not give enough information to decide confidently.

Decision principle:

Prefer "Unclear" over "No" whenever the abstract is silent or ambiguous. A wrong "No" permanently drops a paper, whereas a wrong "Unclear" is resolved during full-text screening.

Special decision rules:

1. A study of only the immediate or acute response to a single exercise session is "No" for Criterion 2.

2. Criterion 3 is "Yes" if there is an exercise-only arm, or a comparison in which one arm differs from another only by exercise. Multi-arm trials that include such an arm alongside other arms (e.g., diet + exercise, usual care) remain eligible; only the exercise-relevant arms are extracted downstream. Exercise inseparably bundled with another intervention (e.g., diet + exercise with no isolating arm) is "No". For a mixed cohort of hypertensive and non-hypertensive participants, the hypertensive participants must also form a defined arm or subgroup; if the paper clearly analyzes the mixed cohort only as a whole, Criterion 3 is "No".

3. Criterion 4 is "No" for observational studies, reviews, commentaries, protocols, and design or methods-only papers.

4. For Criterion 5, statistical support for at least one comparison involving the exercise arm means any of: a p-value, confidence interval, or test statistic; a significance symbol defined in a table legend or footnote; or an explicit statement of significance or non-significance, provided the paper states its significance level.

5. Abstracts often omit population detail, arm structure, and statistics. Mark Criteria 1, 3, and 5 "Unclear" rather than "No" when the abstract is silent on them.

Output:

Copy paper_id into the output. Provide one decision per criterion, in order, as criterion_1 through criterion_5 in the schema. Draw evidence from the title or abstract. Set a criterion's explanation to "NA" when its decision is "Yes"; otherwise give a concise one-sentence explanation.

Prompt for title-and-abstract screening

**Fig C. Prompt for title-and-abstract screening**

Task prompt of the title-and-abstract screening step. Each request contained this prompt, the paper ID, the title and the abstract, with the model’s reasoning effort set to “none”, and the JSON schema of the answer. Shown verbatim.

Task: Step 0b full-text screening.

This is the second pass of paper screening. Decide whether a paper meets the eligibility criteria for a study examining the effect of structured physical exercise training on blood pressure in hypertensive populations. This pass uses the full paper text together with the prior title/abstract screening decisions.

You will receive:

1. paper_id

2. The prior title/abstract screening decisions for criteria 1-5, each given as "Yes" or "Unclear"

3. The full paper markdown

Return a JSON object conforming to the provided output schema.

Relationship to the prior screening:

- The prior title/abstract decisions are context only. The full text is the authoritative source.

- Re-evaluate every criterion against the full text, confirming or revising each prior decision. A prior "Yes" can be overturned (e.g., an abstract may describe a hypertensive population while the analyzed cohort is normotensive), and a prior "Unclear" must be resolved to "Yes" or "No".

Eligibility criteria:

1. Does the study population include individuals with hypertension or elevated blood pressure (prehypertension)?

2. Is a structured exercise training program (multiple sessions over days or weeks) evaluated as an intervention?

3. Can the effect of exercise be isolated from other intervention components (e.g., diet) for the hypertensive population?

4. Is this a completed intervention study with reported results?

5. Is blood pressure (e.g., SBP, DBP, MAP, ambulatory BP) reported as an outcome, with a statistically supported comparison involving the exercise arm?

Decision values:

- "Yes": the full text satisfies the criterion.

- "No": the full text does not satisfy the criterion.

If the full text is ambiguous on a criterion, decide on the reading best supported by the text, and note the ambiguity in that criterion's explanation.

Special decision rules:

1. A study of only the immediate or acute response to a single exercise session is "No" for Criterion 2.

2. Criterion 3 is "Yes" if there is an exercise-only arm, or a comparison in which one arm differs from another only by exercise. Multi-arm trials that include such an arm alongside other arms (e.g., diet + exercise, usual care) remain eligible; only the exercise-relevant arms are extracted downstream. Exercise inseparably bundled with another intervention (e.g., diet + exercise with no isolating arm) is "No". For a mixed cohort of hypertensive and non-hypertensive participants, the hypertensive participants must also form a defined arm or subgroup; if the paper clearly analyzes the mixed cohort only as a whole, Criterion 3 is "No".

3. Criterion 4 is "No" for observational studies, reviews, commentaries, protocols, and design or methods-only papers.

4. For Criterion 5, statistical support for at least one comparison involving the exercise arm means any of: a p-value, confidence interval, or test statistic; a significance symbol defined in a table legend or footnote; or an explicit statement of significance or non-significance, provided the paper states its significance level.

Output:

Copy paper_id into the output. Provide one decision per criterion, in order, as criterion_1 through criterion_5 in the schema. Draw evidence from the paper text. Set a criterion's explanation to "NA" when the full text clearly satisfies it; otherwise give a concise one-sentence explanation.

Prompt for full-text screening

**Fig D. Prompt for full-text screening**

Task prompt of the full-text screening step. Each request contained this prompt, the paper ID, the decisions of the title-and-abstract step as context, the full text of the paper as Markdown, and the JSON schema of the answer; the reasoning effort was “low”. Shown verbatim.

You are a data extraction assistant. You read scientific papers describing physical activity or exercise interventions in hypertensive populations and extract structured information for downstream analysis.

Use only the paper text and/or structured inputs provided by the user. Do not use outside knowledge.

General rules:

1. Capture what the paper says. Do not normalize, classify, compute, convert, or infer values. When the task defines a closed list of options for a field, choose the option directly supported by the paper text; if none fits, use the "other" or "unclear" option the field provides.

2. Every extracted attribute has a status: "reported" (the paper states it), "not_reported" (the paper does not state it), "not_applicable" (the attribute does not apply to this entity), or "unclear" (the paper mentions it but the value cannot be determined). Set value to null unless status is "reported".

3. Every reported value must include the shortest sufficient verbatim evidence quote and a location naming where it was found (for example, "Methods", "Table 2", "Figure 3 legend", "Abstract"). Copy quotes exactly as they appear in the provided text, including LaTeX markup (for example, keep "50\%" and "$t$"). Each evidence field holds one contiguous quote: never join separate passages with "..." and never drop or alter characters. Figures are attached as images after the text, in the order of the "attached_image_k" markers; each marker sits inside its figure block next to the figure caption. A value that appears only in an image may be supported by the text visible in that image, copied exactly, with the location "<caption label> (image)", for example "Figure 2 (image)" (use "attached_image_k (image)" if the caption has no label).

4. When the task provides entity IDs (arm, intervention, time point, outcome), refer to entities only by those IDs and never invent new ones. If something cannot be mapped to a provided ID, describe it in extraction_caveats. When the task asks you to create provisional keys, create them in the requested format, define each key once, and refer only to keys you have defined.

5. If interpretation is uncertain, extract only what is directly supported and explain the ambiguity in extraction_caveats. Do not add a caveat merely because a field is not reported.

6. Evidence quotes should be short. Do not copy entire tables or long blocks unless a field asks for a verbatim passage.

7. Return the requested structured JSON only.

System prompt for information extraction

**Fig E. System prompt for information extraction**

System prompt of the information extraction stage. It was the system message of every request of the skeleton, outcomes, study, interventions, arms and results steps, and of the cross-check and adjudication of the first two. Shown verbatim.

Task: Step 1a-1 skeleton.

Identify the structure of the study: its arms, its interventions, and which arm receives which intervention. Later steps will extract all other details against this skeleton, so focus on identifying each entity correctly and completely. Do not extract demographics, protocol details, outcomes, time points, or results.

You assign provisional keys (A1, A2, …; I1, I2, …) only to express links inside this output. Final IDs are assigned later by code.

First state the design: "parallel" (each participant is allocated or recruited to one arm only), "crossover" (participants receive more than one intervention in sequence), or "other".

1. arms

An arm is a set of participants that the paper allocates or recruits as a distinct group.

- Arms must be disjoint, and together they must cover every participant who was allocated or recruited.

- Never list a group that contains another listed group: no pooled groups such as "all participants" or "all hypertensives", and no subgroups such as "women in the exercise group".

- Include every arm, including comparison populations outside the hypertensive target (for example, a normotensive control group).

- Parallel designs: one arm per allocated group.

- Crossover designs: one arm per randomized sequence (for example, "AB" and "BA") if the paper distinguishes sequences; otherwise one arm containing all participants.

- For each arm give:

- label: the paper's own name for the group.

- appears_as: every exact string under which this arm appears in the text and tables, copied as they appear. This must include every table column header or row label under which the arm's values are reported (for example, "Hypertensive Exercise-Trained ( $\mathrm{N}=11$ )"), plus abbreviations and alternative names. Later checks match table headers against these strings, so a header missing here makes the arm's table values unverifiable.

- n_assigned: number allocated or recruited to the arm, with evidence.

- distinguishing_feature: one short phrase stating what separates this arm from the other arms, with evidence. For a control or comparison arm, state what it lacks or does differently relative to the other arms (for example, "no supervised exercise training"), not what all arms share (for example, advice given to all participants). Describe what the arm consists of or lacks, never just the fact of allocation (not "assigned to the exercise group").

2. interventions

An intervention is a condition that participants receive: an exercise program, a control condition, usual care, no exercise, a sham procedure, etc.

- Distinct protocols are distinct interventions.

- If two arms receive the same protocol (for example, hypertensive and normotensive groups doing the same training), list that protocol once.

- Control conditions are interventions too (for example, "instructed to avoid regular exercise").

- For each intervention give label, appears_as, and distinguishing_feature, as for arms. For an intervention, distinguishing_feature states what the protocol or condition consists of (type, frequency, duration, intensity), not the fact of allocation.

3. arm_interventions

- List every (arm, intervention) pair in which the arm receives the intervention. Every arm must have at least one pair.

- Parallel designs: exactly one pair per arm.

- Crossover designs: one pair per intervention the arm receives, with period_label stating the period or order if the paper defines it; otherwise null.

- Parallel designs: period_label is always null.

Rules:

- Create arms for subgroup analyses (for example, men vs women within an arm) only if the paper allocated or recruited them as separate groups.

- Keep groups that the paper keeps distinct as separate arms.

- List only arms and interventions that the paper describes.

Prompt for the study skeleton

**Fig F. Prompt for the study skeleton**

Task prompt of the skeleton step, which identifies the design, arms, interventions and arm–intervention links. It was given to two independent extractors (OpenAI gpt-6-luna and Google gemini-3.8-flash; medium reasoning effort) together with the paper ID, the Markdown text of the paper, its figures as images and the JSON schema of the answer. Shown verbatim.

Task: Step 1a-2 cross-check of skeleton discrepancies.

You are a reviewer. Two independent extractors identified the study skeleton (arms, interventions, arm-intervention links) for this paper. They disagree on the items listed in the task input. The specification both extractors followed is given in the task input; judge every item by that same specification. For each item you receive:

- item_id

- entity_type (design, arm, intervention, arm_intervention)

- question: what the disagreement is about

- answer_1 and answer_2: the two candidate answers, each with its evidence and location. The order is random and the author of each answer is not disclosed. Either answer, both, or neither may be right.

For each item, re-read the paper and return one verdict:

- "answer_1" or "answer_2": the paper text supports this answer and not the other.

- "neither": the paper text clearly supports a different answer. Give it in corrected_value.

- "cannot_determine": the paper text does not settle the question.

Rules:

- Judge only from the paper text. Do not prefer an answer because of its position, wording, length, level of detail, or confidence. The evidence an answer cites is a claim to check, not a fact.

- Some items ask whether two entries are the same entity (for example, a "Football group" and a "Soccer training group" arm). The two answers are then statements ("one time point" versus "two distinct time points"); judge them like any other pair.

- Some items ask which list of arms or interventions the study has. Judge them by the specification's rules (for example, one arm per randomized sequence only if the paper distinguishes sequences, otherwise one arm for all participants).

- decisive_evidence must be the shortest verbatim quote that settles the question, with its location. A verdict without decisive evidence must be "cannot_determine".

- If both answers are supported (for example, two acceptable labels for the same entity), choose the one whose evidence is more specific and say so in rationale.

- rationale: one or two sentences. Describe the answers by their content (for example, "the 21 allocated, not the 20 who started"), never as "answer_1" or "answer_2", because your rationale may be shown to another reviewer who sees the answers in a different order.

- Return one verdict per item_id, covering every item.

Prompt for cross-checking skeleton discrepancies

**Fig G. Prompt for cross-checking skeleton discrepancies**

Task prompt of the blind cross-check of the skeleton. Both models reviewed every item on which the two skeletons disagreed, saw the two candidate answers in random order without knowing their authors, and received this prompt, the skeleton prompt (Fig F) as the specification, and the paper. Shown verbatim.

Task: Step 1a-3 final adjudication of unresolved skeleton discrepancies.

Two independent extractors identified the study skeleton for this paper and proposed different answers on the items in the task input. Two blind reviewers then re-checked each item against the paper, but they did not agree or could not decide, so the items remain unresolved. The specification all of them followed is given in the task input. For each item you receive:

- item_id, entity_type (design, arm, intervention, arm_intervention), question

- answer_1 and answer_2: the two candidate answers, each with evidence and location. The order is random and the author of each answer is not disclosed.

- critique_1 and critique_2: the two reviewers' verdicts, each with the answer it supports (expressed in your answer_1 / answer_2 labels, or "neither" / "cannot_determine"), its corrected_value (when it supports "neither"), its decisive evidence and location, its rationale, and quote_check, which says whether its decisive evidence was found in the paper text, was read from a figure image, or was not found. The order is random and the reviewer is not disclosed.

You are the final adjudicator. Every item must be decided here; there is no later step. For each item, return one verdict:

- "answer_1" or "answer_2": the answer the paper text better supports. Choose the side you lean towards even when the evidence is not conclusive.

- "neither": the paper text unambiguously supports a different answer. Give it in corrected_value, written in the same form as the candidate answers (for example "21 participants", "Yes, ..." or "No, ...", or the design name). Use this only when the supporting quote leaves no reasonable doubt.

Rules:

- Decide from the paper text. Treat the critiques as arguments to check, not as votes: two critiques that agree can both be wrong, and a critique that cites a quote you cannot find in the paper carries no weight, unless the quote is read from a figure image that you can read yourself.

- decisive_evidence: the shortest verbatim quote that best supports your verdict, with its location. If no passage can be quoted (for example, the answer is an absence), set it to null and say so in the rationale.

- notes: null when the item is clear. When the item is hard (the evidence is not conclusive, the spec is ambiguous, or the answer you did not choose also has merit), write one or two sentences saying why and giving the strongest point for the other answer. A non-null note flags the item for later review, so do not use it for routine remarks.

- Judge every item by the specification in the task input.

- rationale: one or two sentences.

- Return one verdict per item_id, covering every item.

Prompt for adjudicating skeleton discrepancies

**Fig H. Prompt for adjudicating skeleton discrepancies**

Task prompt of the final adjudication of the skeleton items that the two reviewers did not resolve. It was run with Anthropic Claude Sonnet 5.5, which saw the two candidate answers, the verdicts of the two reviewers and the paper. Shown verbatim.

Task: Step 1b-1 outcomes and time points.

Identify the outcome measures of the study, the time points at which they are reported, and which outcome is reported at which time point. Later steps will extract the results of each outcome at each time point, so focus on identifying each outcome and time point correctly and completely: an outcome missed here will have no results, and a spurious one wastes later work. Do not extract arms, interventions, units, measurement methods, or values.

You assign provisional keys (O1, O2, …; T0, T1, …) only to express order and links inside this output. Final IDs are assigned later by code.

1. outcomes

What counts as an outcome:

- A measure assessed to evaluate the effect of, or the response to, the intervention: measured after baseline, or compared over time or between groups.

- The acute response to a single exercise or intervention session (for example the maximum drop in BP after one session) counts as an outcome.

- The paper must report results for it in its own main text, tables, or figures: values, or a stated test result (for example "did not change, P = 0.95"). Include outcomes whose results appear only in a table, only in a figure, or only in text.

- Do not list a measure that is only named or described in Methods without reported results.

- Do not list a measure whose results are only in supplementary materials. A sentence that only points to a supplementary table ("Supplemental Table S6 shows stroke volume") is not a reported result; a sentence in the main text that states the values or the test result is.

- Do not list a measure that the paper attributes only to earlier or other publications, or says is not part of its analysis.

What does not count:

- Baseline-only descriptors such as age, gender, height, or medication use. A characteristic that is also measured after baseline (for example body weight or BMI) is an outcome.

- Adverse events; they are handled separately.

- Process measures: adherence, attendance, dropout, achieved training load, heart rate during training sessions.

- Variables used only as predictors or covariates in a regression or correlation (for example baseline SBP as a predictor of response).

How to list them:

- Use the paper's own name for each outcome.

- Treat measurement variants as distinct outcomes: office SBP, 24-h SBP, daytime SBP, nighttime SBP, central SBP, seated versus standing, and so on, whenever the paper reports them separately.

- Never split one outcome by time point, arm, or subgroup. SBP at week 4 and SBP at week 8 are one outcome; "SBP in men" and "SBP in women" are one outcome. Daytime and nighttime ambulatory SBP are measurement variants, not time points.

- Change scores, percent changes, and group differences of a listed measure are not separate outcomes (for example "Δ SBP" and "% change in SBP" belong to "SBP"). Put their labels in appears_as.

- The same measure expressed in another unit, or divided by an independent size variable such as body weight, height, or body surface area, is one outcome (for example absolute and body-size-normalized values, or two unit systems). Name it by the form the paper reports first, and put the labels of the other forms in appears_as.

- A part expressed as a proportion of the whole it belongs to is a separate outcome from the absolute measure of that part.

- A categorical status derived from a measure is a separate outcome when the paper reports it (for example "proportion with BP < 140/90 mm Hg").

- List derived measures (for example MAP or pulse pressure) separately when the paper reports them.

- Do not list a parent category when its specific measures are listed: not "blood pressure" next to SBP and DBP, not "lipid profile" next to total cholesterol, LDL, HDL, and triglycerides.

- When the paper names the components of a composite but reports results only for the components, list the components, not the composite (for example "sedentary time" and "active time", not "body motion").

- List a parent when the paper reports results for it as its own measure, in addition to its components (for example a global score or total score next to its domains or components).

- For each outcome give:

- outcome_key: O1, O2, … in order of first appearance in Methods, Results, or tables.

- name: the paper's name for the outcome.

- appears_as: every exact string under which this outcome appears, copied as it appears: table row labels, table column headers, figure axis labels, abbreviations, and the labels of its change scores (for example "Clinic systolic BP, mm Hg", "Δ Casual SBP"). Later checks match table rows against these strings, so a row label missing here makes that row's values unverifiable.

- evidence and location for where its results are reported (a result sentence, a table row, or a figure label).

- timepoint_keys: see below.

2. timepoints

List every occasion at which at least one listed outcome has a reported result. An occasion with no reported outcome is not a time point.

- The baseline is T0 with is_baseline true: the pre-intervention occasion that the reported changes are measured against. Order the remaining time points chronologically (T1, T2, …).

- One occasion is one time point even when the paper names it in several ways ("baseline", "pre-training", "PRE", "before training"). Put every name in appears_as.

- A summary over a window or a series of readings (maximum drop, area under the curve, a 24-h mean) is one time point named by its window. List the individual readings of a series as separate time points only if the paper reports results separately for each.

- Do not list protocol milestones at which no outcome is reported (for example "an interval was added at week 6").

- For each time point give:

- label: the paper's own label ("baseline", "week 8", "post-intervention", …).

- appears_as: every exact string under which the time point appears in text, tables, and figures, copied as it appears.

- offset_verbatim: the paper's timing phrase ("after 12 weeks", "4 months", "48 hours after the last session"). Do not convert units.

- evidence and location: the passage that shows the occasion exists and when it is.

3. timepoint_keys of each outcome

Decide the time points in section 2 first; then give each outcome the keys of those time points.

Link only the time points involved in a reported result for the outcome: a value at the time point, a change or comparison that uses it, or a stated test result at it.

- A change or comparison between two occasions (for example "Δ from PRE to POST") links both occasions.

- Link baseline when baseline values, or a change from baseline, are reported for the outcome.

- Do not link a time point only because Methods say the outcome was measured there. If the results reported for the outcome do not involve the time point, leave it out (for example an outcome reported only as a change from PRE to POST is not linked to MID, even if it was also measured at MID).

- Do not link a time point at which the outcome was not reported, even if other outcomes were.

Rules:

- Keep occasions that are at different times as separate time points.

- List only outcomes and time points that the paper describes.

Prompt for outcomes and time points

**Fig I. Prompt for outcomes and time points**

Task prompt of the outcomes step, which identifies the outcomes, the time points and the outcome–time point links. It was given to the same two independent extractors as the skeleton prompt (OpenAI gpt-6-luna and Google gemini-3.8-flash; medium reasoning effort), with the paper ID, the Markdown text of the paper, its figures as images and the JSON schema of the answer. Shown verbatim.

Task: Step 1b-2 cross-check of outcome and time point discrepancies.

You are a reviewer. Two independent extractors identified the outcomes of this study, the time points at which results are reported, and which outcome is reported at which time point. They disagree on the items listed in the task input. The specification both extractors followed is given in the task input; judge every item by that same specification. For each item you receive:

- item_id

- entity_type (outcome, timepoint, outcome_timepoint)

- question: what the disagreement is about

- answer_1 and answer_2: the two candidate answers, each with its evidence and location. The order is random and the author of each answer is not disclosed.

For each item, re-read the paper and return one verdict:

- "answer_1" or "answer_2": the paper text supports this answer and not the other.

- "neither": the paper text clearly supports a different answer. Give it in corrected_value, written in the same form as the candidate answers (for example "Yes, ...", "No, ...", or the Only: form described below).

- "cannot_determine": the paper text does not settle the question.

Rules:

- Judge only from the paper text. Do not prefer an answer because of its position, wording, length, level of detail, or confidence. The evidence an answer cites is a claim to check, not a fact.

- Some items ask whether two entries are the same entity (for example, "SBP" and "Systolic blood pressure" outcomes, or a "baseline" and a "pre-training" time point). The two answers are then statements ("the same" versus "two distinct"); judge them like any other pair.

- Some items ask whether the paper reports a result for an outcome at a time point (entity_type outcome_timepoint). Apply the specification: a value at the time point, a change or comparison that uses it, or a stated test result counts; a Methods sentence saying that the outcome was measured there does not.

- Some items ask whether an entry is a separate outcome or time point of the study. Apply the specification's rules on what counts, including what is excluded.

- Some items ask about a group of outcomes or time points that one extractor lists (for example the rows of one table, or a series of time points). If all of them are separate, or none is, choose the matching answer. If only some are, answer neither and write corrected_value as: Only: "X"; "Y", listing in quotes, as in the question, the members that are separate.

- decisive_evidence must be the shortest verbatim quote that settles the question, with its location. A verdict without decisive evidence must be "cannot_determine".

- If both answers are supported (for example, two acceptable labels for the same entity), choose the one whose evidence is more specific and say so in rationale.

- rationale: one or two sentences. Describe the answers by their content (for example, "the 24-h mean is reported, the individual readings are not"), never as "answer_1" or "answer_2", because your rationale may be shown to another reviewer who sees the answers in a different order.

- Return one verdict per item_id, covering every item.

Prompt for cross-checking outcome and time point discrepancies

**Fig J. Prompt for cross-checking outcome and time point discrepancies**

Task prompt of the blind cross-check of the outcomes and time points. Both models reviewed every item on which the two extractions disagreed, saw the two candidate answers in random order without knowing their authors, and received this prompt, the outcomes prompt (Fig I) as the specification, and the paper. Shown verbatim.

Task: Step 1b-3 final adjudication of unresolved outcome and time point discrepancies.

Two independent extractors identified the outcomes of this study, the time points at which results are reported, and which outcome is reported at which time point. They proposed different answers on the items in the task input. Two blind reviewers then re-checked each item against the paper, but they did not agree or could not decide, so the items remain unresolved. The specification all of them followed is given in the task input. For each item you receive:

- item_id, entity_type (outcome, timepoint, outcome_timepoint), question

- answer_1 and answer_2: the two candidate answers, each with evidence and location. The order is random and the author of each answer is not disclosed.

- critique_1 and critique_2: the two reviewers' verdicts, each with the answer it supports (expressed in your answer_1 / answer_2 labels, or "neither" / "cannot_determine"), its corrected_value (when it supports "neither"), its decisive evidence and location, its rationale, and quote_check, which says whether its decisive evidence was found in the paper text, was read from a figure image, or was not found. The order is random and the reviewer is not disclosed.

You are the final adjudicator. Every item must be decided here; there is no later step. For each item, return one verdict:

- "answer_1" or "answer_2": the answer the paper text better supports. Choose the side you lean towards even when the evidence is not conclusive.

- "neither": the paper text unambiguously supports a different answer. Give it in corrected_value, written in the same form as the candidate answers (for example "Yes, ...", "No, ...", or the Only: form described below). Use this only when the supporting quote leaves no reasonable doubt.

Rules:

- Decide from the paper text. Treat the critiques as arguments to check, not as votes: two critiques that agree can both be wrong, and a critique that cites a quote you cannot find in the paper carries no weight, unless the quote is read from a figure image that you can read yourself.

- Some items ask about a group of outcomes or time points that one extractor lists (for example the rows of one table, or a series of time points). If all of them are separate, or none is, choose the matching answer. If only some are, answer neither and write corrected_value as: Only: "X"; "Y", listing in quotes, as in the question, the members that are separate.

- For outcome_timepoint items, apply the specification: a value at the time point, a change or comparison that uses it, or a stated test result counts as a reported result; a Methods sentence saying that the outcome was measured there does not.

- decisive_evidence: the shortest verbatim quote that best supports your verdict, with its location. If no passage can be quoted (for example, the answer is an absence), set it to null and say so in the rationale.

- notes: null when the item is clear. When the item is hard (the evidence is not conclusive, the specification is ambiguous, or the answer you did not choose also has merit), write one or two sentences saying why and giving the strongest point for the other answer. A non-null note flags the item for later review, so do not use it for routine remarks.

- Judge every item by the specification in the task input.

- rationale: one or two sentences.

- Return one verdict per item_id, covering every item.

Prompt for adjudicating outcome and time point discrepancies

**Fig K. Prompt for adjudicating outcome and time point discrepancies**

Task prompt of the final adjudication of the outcome and time point items that the two reviewers did not resolve. It was run with Anthropic Claude Sonnet 5.5, which saw the two candidate answers, the verdicts of the two reviewers and the paper. Shown verbatim.

Task: Step 1c paper-level facts.

Extract facts that apply to the study as a whole. Do not extract anything that differs between arms (demographics, protocols, results); later steps handle those.

Design:

- design: choose one of parallel_rct, crossover_rct, cluster_rct, factorial_rct, nonrandomized_controlled, single_group_pre_post, other, based on the paper's description.

- washout_verbatim: for crossover designs, the wash-out between periods and any statement on whether period-specific data are reported. not_applicable for other designs.

Population and setting:

- inclusion_criteria_verbatim, exclusion_criteria_verbatim: copy the eligibility criteria.

- countries_verbatim: country or countries where participants were recruited, as stated.

- centers: single or multi.

- n_enrolled_total: number of participants enrolled or screened at the start of the study, before randomization or exclusions. Do not use the number analyzed or followed up; if the paper gives only that, the field is not_reported.

- n_randomized_total: number randomized or allocated across all groups.

- hypertension_definition_verbatim: the criterion or threshold used to define hypertension for eligibility.

Statistics:

- statistical_methods_verbatim: the tests or models used to analyze outcomes.

- significance_threshold_verbatim: the stated alpha (for example, "P<0.05 was considered statistically significant").

Prompt for study-level facts

**Fig L. Prompt for study-level facts**

Task prompt of the study step, which extracts the facts that apply to the study as a whole. It was given to OpenAI gpt-6-luna (medium reasoning effort) with the paper ID, the Markdown text of the paper, its figures as images and the JSON schema of the answer. Shown verbatim.

Task: Step 1d intervention details.

Extract the details of one intervention. The task input gives:

- target_intervention_id

- entity cards for all arms, interventions, and time points in the paper (for context and to avoid confusing interventions)

Return intervention_id equal to target_intervention_id. Extract only information about the target intervention. If a passage describes several interventions, use only the part that applies to the target.

Classification (closed lists; choose what the paper's description directly supports):

- intervention_type: exercise, exercise_plus_other, other_active_non_exercise, stretching_or_low_intensity_control, sham, attention_or_education_control, usual_care, no_intervention, waitlist, other.

- Use exercise_plus_other only when the extra component is part of what is being tested, that is, it is not given identically to the comparison arms (see the entity cards). A component that every arm receives (for example dietary counseling given to all participants) is background: it does not make the type exercise_plus_other (the flags still record it).

- An intervention that consists only of a placebo or sham (a placebo capsule, a low-resistance sham device) has the type sham.

- framed_as_control: true if the paper presents this condition as the control or comparison condition.

Protocol:

- protocol_verbatim: the highest-priority field. Copy the complete Methods description of what participants receiving this intervention did or were instructed to do: frequency, intensity, session structure, modality, total duration, progression, equipment, setting, supervision, and bundled components. For control conditions, copy what participants were told to do or avoid.

Component flags (true or false):

- A flag is true if the paper describes the component as part of what participants of this intervention receive, even if every arm receives it, and false otherwise. A placebo or sham is not a component, so the flag stays false. For a false flag, the evidence is the sentence that shows what the intervention consists of. Use status "not_reported" only if the paper gives no description of this intervention.

- has_exercise: structured exercise or physical activity of any kind that participants perform as part of this intervention. Advice or guidelines to exercise are not exercise: count them under has_behavioral_support. If exercise is applied in every arm over time rather than appearing in the arm label, still set true and explain in extraction_caveats.

- has_diet: a diet intervention.

- has_supplementation: supplement use.

- has_environmental_conditioning: heat therapy, hot water immersion, sauna, altitude, hypoxia, cold exposure, or similar.

- has_behavioral_support: counseling, coaching, education, or behavior-change support.

Program details (for exercise or for any other active intervention such as heat therapy; copy as worded; do not convert "twice weekly" to "2 sessions/week"). Give the prescribed values, not the values achieved by the participants. If several passages apply, separate them with "; ":

- frequency_verbatim

- session_duration_verbatim

- intensity_verbatim: the prescribed intensity and how it was expressed (%HRmax, %HRR, %VO2peak, %1RM, %MVC, RPE, speed, etc.).

- modality_verbatim

- total_duration_verbatim: length of the program.

- progression_verbatim: how frequency, duration, or intensity changed over the program.

Delivery (closed lists):

- supervision: fully_supervised, partly_supervised, unsupervised.

- venue: laboratory_or_clinic, community_or_gym, home, mixed, other.

- delivery_format: individual, group, mixed.

- provider_verbatim: who delivered the intervention.

- equipment_verbatim.

For a no-intervention or usual-care condition, set the program details and delivery fields to not_applicable unless the paper describes them.

Rules:

- Weekly volume is not computed and intensity is not classified.

Prompt for intervention details

**Fig M. Prompt for intervention details**

Task prompt of the interventions step, which was run once per intervention. It was given to OpenAI gpt-6-luna (medium reasoning effort) with the paper, the entity cards of the arms, interventions and time points from the structure step, the ID of the target intervention and the JSON schema of the answer. Shown verbatim.

Task: Step 1e arm details and baseline characteristics.

Extract, for every arm, the arm details and the baseline characteristics of its participants. The task input gives entity cards for all arms, interventions, time points, and outcomes in the paper. Return exactly one record in arms for each arm card, in the order of the cards. A record never holds statistics of another arm: populations are only for subgroups of that same arm.

Part 1: arm details (for each arm)

- stated_bp_status: hypertensive, prehypertensive_or_elevated, normotensive, mixed, not_stated. Base this only on sentences of the text that describe the participants (title, abstract, aim, eligibility and participant description). When the title or aim names the population ("adults with untreated hypertension"), use that label rather than the eligibility range. Never use tables, baseline BP values, or BP-stage counts; those belong to the results. Use mixed only when the text itself names both kinds of participants in the arm. If the text does not describe it, use not_stated with status "not_reported".

- n_completed: number completing the intervention or follow-up.

- medication_status: antihypertensive treatment status of the arm as a whole during the trial. Choose never_treated, untreated_at_entry, stable_during_trial, changed_during_trial, or mixed_within_arm. Use status "not_reported" if the paper does not say.

- One participant or a few who started, stopped, or switched medication do not set the status of the arm; use the paper's description of the arm (for example "untreated hypertension" gives untreated_at_entry) and mention such exceptions in extraction_caveats.

- Use changed_during_trial only when the protocol changes medication for the arm (for example the habitual drug is replaced by placebo for everyone), and mixed_within_arm only when the paper reports that a substantial proportion of the arm differs.

Part 2: baseline characteristics (for each arm)

populations lists the groups of the arm that have baseline statistics. The first entry is the whole arm (subgroup_verbatim null). Add an entry for a subgroup (for example "participants with elevated baseline SBP") only if the paper gives baseline statistics for that subgroup; give the subgroup as the paper names it, and fill that entry only with statistics that describe the subgroup.

Each population has these characteristics, each a list of statistics (an empty list means the paper reports nothing for it):

- age, gender, race_ethnicity, bmi (gender holds the counts or percents of men and women, whether the paper says sex or gender)

- antihypertensive_medications: a list of entries, one per medication category and per individual drug the paper lists for the participants, in the paper's own words (for example "Antihypertensive drugs", "Angiotensin II receptor blockers (ARBs)", "Valsartan"). parent_verbatim is the category the paper lists the entry under (for example "ARBs" for "Valsartan"), null for a top-level entry. Do not invent categories and do not sort drugs into classes.

- comorbidities: a list of entries, one per condition the paper gives for the participants (condition_verbatim as the paper names it, for example "type 2 diabetes", "metabolic syndrome").

Look in the whole paper for each characteristic: tables, Results, Methods, Abstract, Discussion, figure legends, and figures. Do not skip a characteristic because the paper does not say when it was measured or because it appears only in the Abstract or Discussion; if it describes the participants of the arm before the intervention, or the sample as a whole, it is a statistic. Descriptions count: "middle-aged women" or "approximately 45 years old" are statistics of kind text.

A statistic is one reported value:

- arm_ids: every arm the reported value covers, always including the arm you are filling in. A pooled value ("All participants (n=25)", "women (n=25)" for two arms) is listed under each arm it covers, with the same list of arm IDs. Never copy a value of one arm to another arm.

- category_label: for a count or percent in gender and race_ethnicity, the category that is counted (for example "women"). When category names repeat under different headers, start the label with the header: "Ethnicity: Decline to answer", "Race: White". Null otherwise (a medication or comorbidity entry already names what is counted).

- kind: mean, median, count, percent, range, min, max, text.

- value: the number as printed, or null for text. value_text: the verbatim text for kind text or for a value that is not a single number (for example "13/7", "middle-aged"); otherwise null.

- spread_type: sd, se, ci, iqr, range, none, unclear. Use the convention stated for the table (for example "Values are mean ± SE") or stated generally in the paper. If no convention is stated for a "±" value, use unclear. Give spread for an SD or SE, and low and high (with level_verbatim, for example "95% CI") for an interval or range.

- unit_verbatim: unit as printed, or null.

- n: the number of participants the statistic is based on, only if the paper prints it for this statistic.

- Give a count and its percentage ("26 (72%)") as one count; give a percent statistic only when no count is printed.

Sources:

- sources lists every place the statistic comes from. Each entry has kind (table, text, figure_legend, footnote), location (for example "Table 1", "Abstract", "Results, 3.1"), evidence (one contiguous quote copied exactly, including LaTeX markup), and table_cell.

- The same value reported in Table 1, in the Results text, and in the Abstract is one statistic with three sources. If the values differ (rounding, a different analysis), make separate statistics and explain the difference in extraction_caveats.

- A table cell is a source of kind table: evidence is the cell text (or the row), and table_cell gives the table location, the row header path, and the column header path that locate the cell (copy header text exactly, outermost to innermost). For text, figure legends, and footnotes, table_cell is null. If a value comes from a figure image, say so in location (for example "Figure 1 (image)").

Rules:

- Baseline values of an outcome listed in the outcome cards (for example baseline SBP when SBP is an outcome) belong to results and are not extracted. The exception is BMI: its baseline value is always extracted, even when BMI is an outcome.

- Age groups, BMI categories, and hypertension stage are not classified.

Prompt for arm details and baseline characteristics

**Fig N. Prompt for arm details and baseline characteristics**

Task prompt of the arms step, which extracts the details and the baseline characteristics of every arm. It was given to OpenAI gpt-6-luna (medium reasoning effort) with the paper, the entity cards of the arms, interventions, time points and outcomes, and the JSON schema of the answer. Shown verbatim.

Task: Step 1f results for one outcome.

Extract every comparison reported for one outcome, across all arms, interventions, and time points. The task input gives:

- target_outcome_id and its outcome card

- entity cards for all arms, interventions, arm-intervention links, and time points

- table_notes: footnotes and captions parsed from the paper's tables and figures, keyed by location and symbol

Return outcome_id equal to target_outcome_id.

Each result row is one comparison between side_1 and side_2. It holds the value the paper reports for each side, the reported difference between the sides, and the p-value of the comparison. Every row has two sides, and the two sides must differ in their arms, interventions, or time points.

A side is a group of participants at a time point:

- arm_ids, intervention_ids, timepoint_ids: the IDs that make up the side, each as a list. A side that pools several arms, interventions, or time points lists all of them (for example arm_ids ["a1", "a2"] for two pooled arms, intervention_ids ["i1", "i2"] for a result pooled over two interventions). Never leave a list empty. Pool only what the paper pools.

- reference_timepoint_id: null for a value at the time point. It is set only when the row compares the changes of two groups (see below): then each side is the change from the reference time point to timepoint_ids, and the reference is set on both sides with the same value.

- n_analyzed: the number of participants analyzed for this side, only if the paper prints it for this result (in the cell, the column header, a table note, or the sentence); otherwise null. Do not copy group sizes from other parts of the paper.

- stat: the value the paper reports for the side, or null if it does not report one. kind is mean, median, count, or percent (a side with a reference time point holds the mean, median, count, or percent change from that reference). spread_type is sd, se, ci, iqr, range, none, or unclear; give spread for an SD or SE, and low and high (with level_verbatim, for example "95% CI") for an interval or range.

The comparisons (side_1 minus side_2, as the paper prints it; the kind of comparison follows from the sides, do not label it):

- A group over time: side_1 is the group at the later time point, side_2 the same group at the earlier time point, both without a reference. contrast is the reported difference (the change).

- A change reported as a number for one group (for example "Δ −7.4, p=.003" in one group, or a within-group change column): this is a group over time. side_1 is the group at the later time point, side_2 the same group at the earlier time point, reference null on both sides, side stats null unless the values at the two time points are also reported; the change goes in contrast. Never put a change in a side of a within-group row, and never pair a time point with itself.

- Two groups at one time point: side_1 is the exercise or more active group, side_2 the comparator.

- Two groups in their change over time (for example a time×group interaction): each side is a group's change (timepoint_ids = end time point, reference_timepoint_id = start time point), with the group's change in stat if reported. contrast is the difference between the two changes, if reported.

- Two interventions in the same arm (crossover): side_1 and side_2 have the same arm_ids and different intervention_ids.

- A pooled effect (for example the effect of training pooled over a placebo and a medication trial): list the pooled IDs on both sides. Training pooled over two trials compares the same pooled group at two time points, so both sides have intervention_ids ["i1", "i2"] and the time points differ. A medication effect pooled over time compares two interventions at the same pooled time points, so the time points are the same on both sides and the interventions differ. Use the sides, not the author's wording ("main effect"), to decide.

- Both sides of a time comparison list the same interventions. In a design with a training program between two time points and separate test conditions at each time point (for example placebo and medication trials), list the test conditions on the sides and do not add the training program itself.

Other row fields:

- contrast: the difference the paper reports between side_1 and side_2, as printed, or null. kind is mean_difference, median_difference, relative_difference_percent (a difference expressed as a percent of the comparator or earlier value), or other_effect (ratios, odds ratios, effect sizes). Give the author's direction in direction_verbatim when the wording gives one ("in favor of EX").

- effect_label_verbatim: the paper's own name for the comparison ("PRE vs POST", "time×group interaction", "main effect of training", "Δ"), or null.

- subgroup_verbatim: the subgroup of the arm(s) that the comparison is restricted to, as the paper names it ("participants with elevated baseline SBP", "women", "medication-naïve"), or null when the comparison uses the whole arm(s). Use the same wording for the same subgroup in every row, and never mix subgroup values and whole-arm values in one row. A comparison between two subgroups of the same arm (women vs men, medicated vs not medicated) cannot be written with two sides of the same arm: record it in other_statistics.

- analysis_variant: primary unless the paper reports several analyses of the same result (itt, per_protocol, completers, adjusted, unadjusted, other).

- direction_verbatim: the author's wording of the direction or of a result described without numbers ("decreased significantly in the trained group", "in favor of EX").

What becomes a row:

- Extract every comparison the paper reports for the target outcome, including comparisons with other arms (for example a normotensive group), pooled effects, and interaction terms that compare groups. If you are unsure whether a comparison matters, extract it and record its name in effect_label_verbatim.

- Every value reported for the outcome must appear on a row. Make a time-comparison row for a group only when the paper reports something for it: a value at either time point, a difference, a p-value, a significance statement, or the author's wording of the direction. Never output a row that has none of these. Make a between-group row when the paper reports a difference, a p-value, or a statement for it.

- Report each comparison once. If the same numbers appear in several places (abstract, text, table, figure legend, footnote), keep one row and list every place in sources. If the numbers differ (a different analysis), keep separate rows and explain the difference in extraction_caveats.

- Figure-only results: if the figure legend or text gives numbers, extract them. Otherwise add a row with null stats and the author's wording in direction_verbatim or significance.

p-values and significance:

- p_value_verbatim: exactly as printed ("P=0.03", "P<0.05", "NS"); null if none is printed.

- A sentence can contain several p-values. Attach each p-value to the statistic it is labeled with or directly follows. A p-value labeled "P interaction", "time×group", "between-group", or similar belongs to the comparison between groups; a p-value in parentheses right after a within-group change belongs to that change. Never use a p-value given for another outcome or another statistic.

- p_method_verbatim: the test or model behind the p-value as the paper names it ("two-way repeated-measures ANOVA", "paired t-test", "Bonferroni post-hoc"), or null if the paper does not say.

- significance: use it only when the paper says a comparison is significant or not significant (or a tendency) and prints no p-value for it, for example through a footnote symbol. If a p-value is printed for the comparison, leave significance null. claim is significant, not_significant, or tendency; wording_verbatim is the author's wording. Do not look for the significance level elsewhere in the paper and do not write it as a p-value. If the note itself states a level ("* P<0.05"), put it in p_value_verbatim as printed.

Sources:

- sources lists every place the row comes from. Each entry has kind (table, text, figure_legend, footnote), location (for example "Abstract", "Table 2", "Results, 3.3"), evidence (one contiguous quote copied exactly, including LaTeX markup), and supports: which parts of the row it supports (side_1, side_2, contrast, p_value, significance).

- A table cell is a source of kind table: evidence is the cell text, and table_cell gives the table location and the row and column header paths that locate the cell. A table caption or table note, including a footnote that gives the meaning of a symbol, is a source of kind footnote. A source of kind footnote, text, or figure_legend has table_cell null.

- List only places where the numbers, the p-value, or the significance statement appear. Do not list passages that only describe the statistical method or the data format (for example "Data are presented as mean ± SD" or the Statistical analysis section).

Locating table cells (table_cell):

- row_header_path lists row labels from outermost to innermost. A row that spans the full table width (for example, \multicolumn{4}{|l|}{Clinic systolic BP, mm Hg}) is a parent label for the rows beneath it, until the next such row. A \multirow label applies to every row it spans.

- column_header_path lists column headers from the top header level to the bottom. A \multicolumn header applies to every column it spans.

- Copy header text exactly as it appears, including LaTeX markup and symbols.

Rules for tables:

- One cell can feed several rows. For example, "$130 \pm 5 \dagger^{*}$" in the post-intervention row for arm a1 gives: (1) a row comparing a1 at post (side_1, value 130, SD 5) with a1 at pre (side_2, value from the pre cell); if the table notes say † means significantly lower than pre-training, add significance {significant, "Significantly lower than pre-training"} to that row, with the post cell, the pre cell, and the † footnote line as sources; (2) a row comparing a1 at post with the comparator named in the * note (for example the normotensive arm at post), with its own significance or p-value.

- When a symbol marks a comparison, side_1 is the arm whose cell carries the symbol, and side_2 is the comparator named in the note.

- A row produced from a symbol points to the cell that carries the symbol.

- A cell without a symbol carries no p-value or significance from the notes.

- Use the table notes to read symbols and dispersion. If a note says "Values are mean ± SE", every ± value from that table has spread_type "se".

- A column for a pooled group, a difference, or p-values is not an arm: report differences and p-values as comparison rows between the arms they refer to.

other_statistics: record statistics about the target outcome that are not a comparison between two sides (for example, a correlation between change in this outcome and another variable) verbatim, with location. Each entry must be a complete statement that names what is compared or related and gives the statistic, in the paper's words. Do not record bare table cells or the rows of a regression or correlation table. A pooled main effect or an interaction that compares two defined sides is a result row, not an other_statistics entry.

Rules:

- Results of any other outcome, even from the same table, are not extracted.

- Numbers, differences, intervals, and p-values are copied as printed, keeping the sign; none is computed, converted, or estimated.

Prompt for the results of one outcome

**Fig O. Prompt for the results of one outcome**

Task prompt of the results step, which was run once per outcome. It was given to OpenAI gpt-6-luna (medium reasoning effort) with the paper, the entity cards of the arms, interventions, links and time points, the ID of the target outcome, the table footnotes and captions parsed from the paper, and the JSON schema of the answer. Shown verbatim.

**Table A. PubMed search query and filters**

| **Group** | **Terms (combined with OR)** |
| --- | --- |
| Hypertension | 'hypertension' OR 'high blood pressure' OR 'elevated blood pressure' OR 'prehypertension' OR 'pre-hypertension' OR 'essential hypertension' OR 'primary hypertension' OR 'resistant hypertension' |
| Exercise or control condition | 'physical exercise' OR 'exercise' OR 'physical activity' OR 'exercise training' OR 'exercise program' OR 'exercise protocol' OR 'aerobic exercise' OR 'endurance exercise' OR 'resistance exercise' OR 'strength training' OR 'interval training' OR 'isometric' OR 'sedentary control' OR 'inactive control' OR 'usual care' |
| Blood pressure | 'blood pressure' OR 'arterial pressure' OR 'systolic blood pressure' OR 'diastolic blood pressure' OR 'ambulatory blood pressure' |
| Filters | Published 2005/01/01–2025/12/31; abstract and full text available; randomized controlled trial; English; humans; adults (≥19 years, PubMed age-group definitions) |

The three groups of terms were combined with AND. Records were retrieved from PubMed (Methods; Fig A).

**Table B. LLM calls, cost and runtime per step**

| **Step** | **Model** | **Calls** | **Mean input tokens** | **Mean output tokens** | **Mean cost per call (US$)** | **Total cost (US$)** | **Mean runtime per call (s)** |
| --- | --- | --- | --- | --- | --- | --- | --- |
| **Screening** | | | | | | | |
| Title and abstract | gpt-6-luna | 2,548 | 2,113 | 283 | 0.0004 | 0.90 | 2.6 |
| Full text | gpt-6-luna | 453 | 14,655 | 347 | 0.0016 | 0.74 | 3.3 |
| **Information extraction** | | | | | | | |
| Skeleton | gpt-6-luna | 255 | 19,204 | 2,139 | 0.0030 | 0.76 | 16.2 |
| Skeleton | gemini-3.8-flash | 255 | 18,872 | 3,518 | 0.0273 | 6.97 | 22.4 |
| Skeleton cross-check | gpt-6-luna | 23 | 21,603 | 706 | 0.0025 | 0.06 | 8.8 |
| Skeleton cross-check | gemini-3.8-flash | 23 | 20,886 | 3,542 | 0.0289 | 0.67 | 27.4 |
| Skeleton adjudication | Claude Sonnet 5.5 | 16 | 28,734 | 429 | 0.0618 | 0.99 | 4.1 |
| Outcomes | gpt-6-luna | 255 | 19,820 | 4,276 | 0.0041 | 1.05 | 32.9 |
| Outcomes | gemini-3.8-flash | 255 | 19,710 | 9,141 | 0.0490 | 12.50 | 56.3 |
| Outcomes cross-check | gpt-6-luna | 115 | 22,368 | 1,398 | 0.0029 | 0.34 | 18.1 |
| Outcomes cross-check | gemini-3.8-flash | 115 | 22,082 | 5,116 | 0.0357 | 4.11 | 35.2 |
| Outcomes adjudication | Claude Sonnet 5.5 | 57 | 35,052 | 496 | 0.0751 | 4.28 | 4.6 |
| Study | gpt-6-luna | 255 | 19,076 | 2,216 | 0.0030 | 0.77 | 20.4 |
| Interventions | gpt-6-luna | 624 | 21,560 | 2,721 | 0.0035 | 2.18 | 25.7 |
| Arms | gpt-6-luna | 255 | 23,298 | 5,633 | 0.0050 | 1.29 | 38.6 |
| Results | gpt-6-luna | 4,679 | 24,490 | 4,176 | 0.0045 | 21.03 | 30.1 |
| **Total** |  | **10,183** |  |  |  | **58.63** |  |

Calls are those of one run of the pipeline: 112 repeated calls (retries after a truncated or unparsable answer, and reruns) are left out, and the costs are those of the calls counted. Cost was computed from the token counts that the pipeline logged for every call, with standard-tier list prices on 3 November 2026, in US$ per million input, cached input and output tokens: gpt-6-luna 0.10, 0.01 and 0.50; gemini-3.8-flash 0.75, 0.075 and 3.75 (price through 31 December 2026); Claude Sonnet 5.5 2.00, 0.10 and 10.00. Input tokens include the attached figures, and output tokens include reasoning tokens. Screening cost US$1.64 and information extraction US$56.99. The runtime of a call is the time from the completion of the previous call to the completion of this one, in runs that sent one call at a time, so it includes local processing; 7 intervals that spanned a break between runs were removed as outliers (above the third quartile plus three interquartile ranges, with an output rate below one third of the step's median). The main run of the results step sent calls in parallel, so its runtime comes from a sequential run of 199 calls that filled in missing outcomes. Calls for different papers are independent and can run in parallel, so the total time depends on the degree of parallelism and is not reported. LLM, large language model.
